# Temporal dynamics and drivers of durable HIV viral load suppression and persistent high- and low-level viremia during Universal Test and Treat scale-up in Uganda: a population-based study

**DOI:** 10.1101/2023.06.15.23291445

**Authors:** Joseph G. Rosen, Robert Ssekubugu, Larry W. Chang, Victor Ssempijja, Ronald M. Galiwango, Joseph Ssekasanvu, Anthony Ndyanabo, Alice Kisakye, Gertrude Nakigozi, Katherine B. Rucinski, Eshan U. Patel, Caitlin E. Kennedy, Fred Nalugoda, Godfrey Kigozi, Oliver Ratmann, Lisa J. Nelson, Lisa A. Mills, Donna Kabatesi, Aaron A.R. Tobian, Thomas C. Quinn, Joseph Kagaayi, Steven J. Reynolds, M. Kate Grabowski

**Affiliations:** Department of International Health, Johns Hopkins Bloomberg School of Public Health, Baltimore, Maryland, United States of America; Rakai Health Sciences Program, Entebbe, Uganda; Division of Infectious Diseases, Johns Hopkins School of Medicine, Baltimore, Maryland, United States of America; Department of Epidemiology, Johns Hopkins Bloomberg School of Public Health, Baltimore, Maryland, United States of America; Clinical Monitoring Research Program Directorate, Frederick National Laboratory for Cancer Research, Frederick, Maryland, United States of America; Department of Mathematics, Imperial College, London, United Kingdom; Division of Global HIV and TB, Centers for Disease Control and Prevention, Kampala, Uganda; Division of Pathology, Johns Hopkins School of Medicine, Baltimore, Maryland, United States of America; Division of Intramural Research, National Institute of Allergy and Infectious Diseases, National Institutes of Health, Bethesda, Maryland, United States of America

**Keywords:** HIV treatment, HIV viremia, antiretroviral therapy, Treat All, prospective cohort, sub-Saharan Africa

## Abstract

**Introduction:** Population-level data on durable HIV viral load suppression (VLS) following implementation of Universal Test and Treat (UTT) in Africa are limited. We assessed trends in durable VLS and viremia among persons living with HIV in 40 Ugandan communities during UTT scale-up.

**Methods:** In 2015-2020, we measured VLS (defined as <200 RNA copies/mL) among participants in the Rakai Community Cohort Study, a longitudinal population-based HIV surveillance cohort in southern Uganda. Persons with unsuppressed viral loads were characterized as having low-level (200-999 copies/mL) or high-level (≥1,000 copies/mL) viremia. Individual virologic outcomes were assessed over two consecutive RCCS survey visits (i.e., visit-pairs; ∼18 month visit intervals) and classified as durable VLS (<200 copies/mL at both visits), new/renewed VLS (<200 copies/mL at follow-up only), viral rebound (<200 copies/mL at initial visit only), or persistent viremia (<200 copies/mL at neither visit). Population prevalence of each outcome was assessed over calendar time. Community-level prevalence and individual-level predictors of persistent high-level viremia were also assessed using multivariable Poisson regression with generalized estimating equations.

**Results:** Overall, 3,080 participants contributed 4,604 visit-pairs over three survey rounds. Most visit-pairs (72.4%) exhibited durable VLS, with few (2.5%) experiencing viral rebound. Among those with viremia at the initial visit (*n*=1,083), 46.9% maintained viremia through follow-up, 91.3% of which was high-level viremia. One-fifth (20.8%) of visit-pairs exhibiting persistent high-level viremia self-reported antiretroviral therapy (ART) use for ≥12 months. Prevalence of persistent high-level viremia varied substantially across communities and was significantly elevated among young persons aged 15-29 years (versus 40-49-year-olds; adjusted risk ratio [adjRR]=2.96; 95% confidence interval [95%CI]:2.21-3.96), men (versus women; adjRR=2.40, 95%CI:1.87-3.07), persons reporting inconsistent condom use with non-marital/casual partners (versus persons with marital/permanent partners only; adjRR=1.38, 95%CI:1.10-1.74), and persons exhibiting hazardous alcohol use (adjRR=1.09, 95%CI:1.03-1.16). The prevalence of persistent high-level viremia was highest among men <30 years (32.0%).

**Conclusions:** Following universal ART provision, most persons living with HIV in south-central Uganda are durably suppressed. Among persons exhibiting viremia, nearly half maintain high-level viremia for ≥12 months and report higher-risk behaviors associated with onward HIV transmission. Enhanced linkage to HIV care and optimized treatment retention could accelerate momentum towards HIV epidemic control.

## Introduction

Universal Test and Treat (UTT) signaled a global paradigm shift in HIV control efforts through expanded antiretroviral therapy (ART) eligibility. Randomized trials in sub-Saharan Africa demonstrated population-level benefits of UTT implementation, with increased population viral load suppression (VLS) preceding significant HIV incidence declines.^1–4^ UTT strategies have rapidly expanded treatment coverage in high-burden settings like Uganda, where the proportion of persons living with HIV on ART increased four-fold over the last decade.^5^ In some areas, ART use is approaching or has exceeded the ambitious 95-95-95 targets for HIV elimination.^6^

Geographic and demographic disparities in treatment outcomes throughout Africa, nevertheless, suggest person-level and place-based factors can modify HIV care engagement and ART adherence, attenuating the effectiveness of evidenced-based epidemic control strategies like UTT. Four seminal UTT trials in sub-Saharan Africa reported suboptimal ART linkage in specific populations (i.e., men, youth, migrants), and none of the UTT arms achieved globally established targets for population VLS (≥90%).^1–3,7^ These findings suggest that UTT implementation alone may be insufficient to closing gaps along the HIV care continuum, especially for populations that are currently underserved by the existing landscape of HIV services.^8,9^

Furthermore, available evidence on longitudinal virologic outcomes is derived primarily from clinically engaged populations, who may be fundamentally incomparable to care-disengaged or treatment-inexperienced persons. Population-based estimates offer a more comprehensive assessment of progress towards HIV epidemic control, but population-level studies examining durable VLS or sustained VLS in individuals are limited.^10–12^ Only one study has been conducted in a generalized HIV epidemic setting and was implemented in four hyperendemic Lake Victoria fishing communities (HIV prevalence: 38-43%),^12^ with demographic and sexual behavior profiles atypical of most African communities.^13,14^ This study observed moderate levels of persistent viremia amidst large increases in durable VLS but did not distinguish between high-(≥1,000 HIV RNA copies/mL) and low-level (200-999 copies/mL) viremia,^12^ which has been linked to subsequent virologic failure and HIV-1 drug resistance.^15–17^

Building from our prior work characterizing population-level viremia during early UTT rollout in fishing communities,^12^ we examined longitudinal patterns and factors associated with durable VLS following mass scale-up of UTT in 40 communities in southern Uganda, including 36 rural agrarian and semi-urban trading communities and four Lake Victoria fishing communities. Our analyses offer unique insights into the evolving dynamics of population-level HIV viremia during a period of substantial HIV service expansion and UTT rollout, including the relative fractions of high- and low-level viremia, and the characteristics of sub-populations who maintain viremia despite universal ART eligibility.

## Methods

### Study design

Data were derived from the Rakai Community Cohort Study (RCCS)—an open, longitudinal population-based HIV surveillance cohort implemented across 40 communities in and around Rakai, Uganda.^13^ Located in south-central Uganda, the RCCS study area is characterized by a heterogeneous HIV epidemic, with high HIV burdens varying substantially in magnitude across communities (range: 9-43%).^13^

Households in enumerated RCCS communities are censused biennially (every ∼12-18 months), and individuals aged 15-49 years residing in RCCS catchment areas for ≥6 months (≥1 months in fishing communities) are eligible for RCCS participation. Enumerators administer structured surveys to RCCS participants measuring household characteristics, sexual behaviors, and HIV service utilization. HIV status is determined using a validated algorithm of three rapid tests in the field and confirmatory testing by enzyme immunoassay in the laboratory.^18^ Viral load testing on stored plasma of participants with confirmed HIV infection is performed using the Abbott RealTi*m*e HIV-1 assay (Abbott Molecular, Inc., Des Plaines, IL).

For this study, inclusion was restricted to persons living with HIV contributing at least two study visits over three survey rounds: Round 17 (February 2015 to September 2016), Round 18 (October 2016 to May 2018), and Round 19 (June 2018 to October 2020). The observation period coincided with various shifts in HIV service delivery in Uganda, most notably UTT scale-up, which began in the four Lake Victoria fishing communities in 2014, then in inland communities in December 2016.^12^

### Viral load suppression measures

The unit of analysis was a visit-pair, defined as two consecutive study visits (V*_i_* + V*_i+j_*) during the observation period. Participants contributed up to two visit-pairs to the analysis. Individuals with a missed visit at Round 18 contributed one visit-pair to the analysis, in which their Round 17 data served as their index visit (V*_i_*) and their Round 19 data as their follow-up visit (V*_i+j_*).

Although low-level viremia is defined in some contexts as any detectable viral load >50 copies/mL,^15,19^ a VLS cutpoint of <200 HIV RNA copies/mL was selected for the present study to minimize statistical “noise” around the assay’s lower limit of detection and distinguish unsuppressed persons with transmissible viremia from those exhibiting transient, low-risk viremic “blips”.^17,20,21^ Four longitudinal virologic outcomes were identified within visit-pairs: (1) durable VLS (<200 copies/mL across visits), (2) new/renewed VLS (<200 copies/mL at follow-up only), (3) viral rebound (<200 copies/mL at index visit only), and (4) persistent viremia (<200 copies/mL at neither visit). Viremia was stratified by two categories based on viral copy counts, per World Health Organization guidelines: low-level viremia (200-999 copies/mL) and high-level viremia (≥1,000 copies/mL).^22^

### Statistical analysis

Data were managed and analyzed in Stata/IC 15.1 (StataCorp LLC, College Station, TX). First, the characteristics of individuals included in the analytic cohort and those excluded from analysis (i.e., lost to follow-up) were compared using Pearson’s χ^2^ (for categorical variables) and Wilcoxon rank-sum tests (for continuous variables) (**Table S1, Figure S1**). Probabilities of study inclusion were then estimated from a multivariable binomial logistic regression model of factors associated with lost to follow-up (**Table S2**). Stabilized inverse probability selection weights were then constructed from these adjusted marginal probabilities (**Table S3**) and applied to subsequent descriptive analyses to mitigate potential attrition bias induced through exclusion of participants with fewer than two study visits.

Unweighted and weighted prevalence estimates for visit-pair-level virologic outcomes were then calculated and compared across age groups, gender, and community types. Sensitivity analyses employing alternative VLS cutpoints (<50, <400, <1,000 copies/mL) were conducted to examine whether prevalence estimates of longitudinal virologic outcomes varied across VLS thresholds (**Table S4**). Among persons contributing three study visits to the analysis, prevalence estimates for longitudinal virologic outcomes were measured to examine 5-year viral load trajectories during UTT scale-up.

Next, within each visit-pair, the probabilities of high-level and low-level viremia at follow-up were assessed conditional on virologic outcomes at the index visit. This analysis was then repeated for each calendar period (index survey visit: February 2015 to September 2016 or October 2016 to May 2018) (**Figure S2**) and community type (**Figure S3**). Among visit-pairs exhibiting persistent high-level viremia, the fraction of persons self-reporting ART use for ≥12 calendar months (i.e., self-reported current ART use at both index and follow-up visits) was calculated (**Table S5**), and correlates of persistent high-level viremia versus durable VLS were identified among persons on ART for ≥12 months using Pearson’s χ^2^ and Wilcoxon rank-sum tests (**Table S6**).

To further evaluate temporal patterns in persistent high-level viremia during UTT scale-up, prevalence estimates for visit-pair-level virologic outcomes were compared over calendar period. Regional- and community-level prevalence estimates for persistent high-level viremia were ascertained by aggregating the cumulative proportion of visit-pairs exhibiting persistent viremia within 10 regions (consisting of two to eight communities) and within each of the 40 communities. Non-parametric Mann-Whitney U tests were then used to examine whether the prevalence of persistent high-level viremia differed significantly (*p*<0.05) over calendar period within individual regions (**Table S7**) and communities (**Table S8**), respectively. Gender-specific prevalence estimates of persistent high-level viremia were also calculated and compared over calendar period within individual regions (**Table S7**).

Lastly, multivariable Poisson regression—overall and stratified by gender and community type, respectively—with generalized estimating equations, exchangeable covariance matrices, and robust standard errors modeled individual-level factors associated with persistent high-level viremia, relative to sustained or new/renewed low-level viremia or suppression (<1,000 copies/mL at both visits or follow-up visit only). Assessed predictors included theorized demographic (age, gender, marital status, education, household wealth, migration, community of residence) and behavioral factors (number of past-year sexual partnerships, condom use consistency across sexual partner types, transactional sex, hazardous alcohol use, and intimate partner violence perpetration or victimization).^12,23–25^ **Table S1** summarizes variable definitions for each factor. Within each visit-pair, demographic factors were derived from a participant’s index visit (V*_i_*), while behavioral factors were derived from the follow-up visit (V*_i+j_*).

Finally, stabilized inverse probability of censoring weights were calculated to account for differential inclusion of participants contributing three visits (or two visit-pairs) to the analysis relative to individuals contributing only two visits (or one visit-pair) (**Table S9**). Inverse probability of selection and censoring weights were then multiplied together to calculate a final analytic weight for multivariable models (**Figure S4**). An optimal working correlation structure was identified through quasilikelihood under the independence model criterion (QIC) values (**Table S10**).^26^ Model-wise deletion was used to handle records with missing observations, which accounted for <1% of all visit-pairs.

### Ethics

The Uganda Virus Research Institute Research and Ethics Committee and the Johns Hopkins University School of Medicine Institutional Review Board reviewed and approved the study protocol. Adults (≥18 years) and emancipated minors provided written informed consent prior to study procedures. Written assent and parental consent were obtained for unemancipated minors aged 15-17 years.

## Results

From February 2015 to October 2020, 33,219 individuals participated in the RCCS, of whom 17.5% (*n* = 5,814) were persons living with HIV (**Figure S1**). Persons whose index visits occurred at Round 19 (15.7%, *n* = 913) were ineligible and, therefore, excluded. Of the remaining 4,901 participants eligible for study inclusion, 3,080 (62.8%) had at least two study visits during the observation period and contributed 4,604 visit-pairs to the analysis. The remaining 1,821 individuals, largely comprised of out-migrants (60.9%), were excluded.

**Table S2** compares index survey visit characteristics of individuals included in the analytic cohort (median age: 34 years, 61.9% women) to those who were excluded (median age: 31 years, 63.7% women). Excluded individuals were significantly (*p*<0.05) younger, wealthier, more mobile, and more likely to exhibit HIV-related risk behaviors (e.g., more sexual partners, hazardous alcohol use) relative to the analytic cohort. The analytic cohort also had significantly better HIV outcomes compared to the excluded population, including self-reported history of ART use (69.8% vs. 57.4%, *p*<0.001) and VLS (71.1% vs. 59.3%, *p*<0.001).

### Prevalence of longitudinal viral load outcomes

**Table 1** presents unweighted and weighted prevalence of longitudinal virologic outcomes at visit-pair level. Most visit-pairs exhibited durable VLS (72.4%) or achieved VLS at the subsequent follow-up visit (13.3%). The prevalence of persistent viremia was 11.8%, and few visit-pairs exhibited viral rebound (2.5%). Weighted estimates did not differ substantially from unweighted estimates. Prevalence estimates of longitudinal virologic outcomes remained stable when higher VLS cutpoints were implemented, but the overall fraction of durable VLS declined substantially when a VLS cutpoint of <50 copies/mL was used (**Table S4**).

**Table 1.**
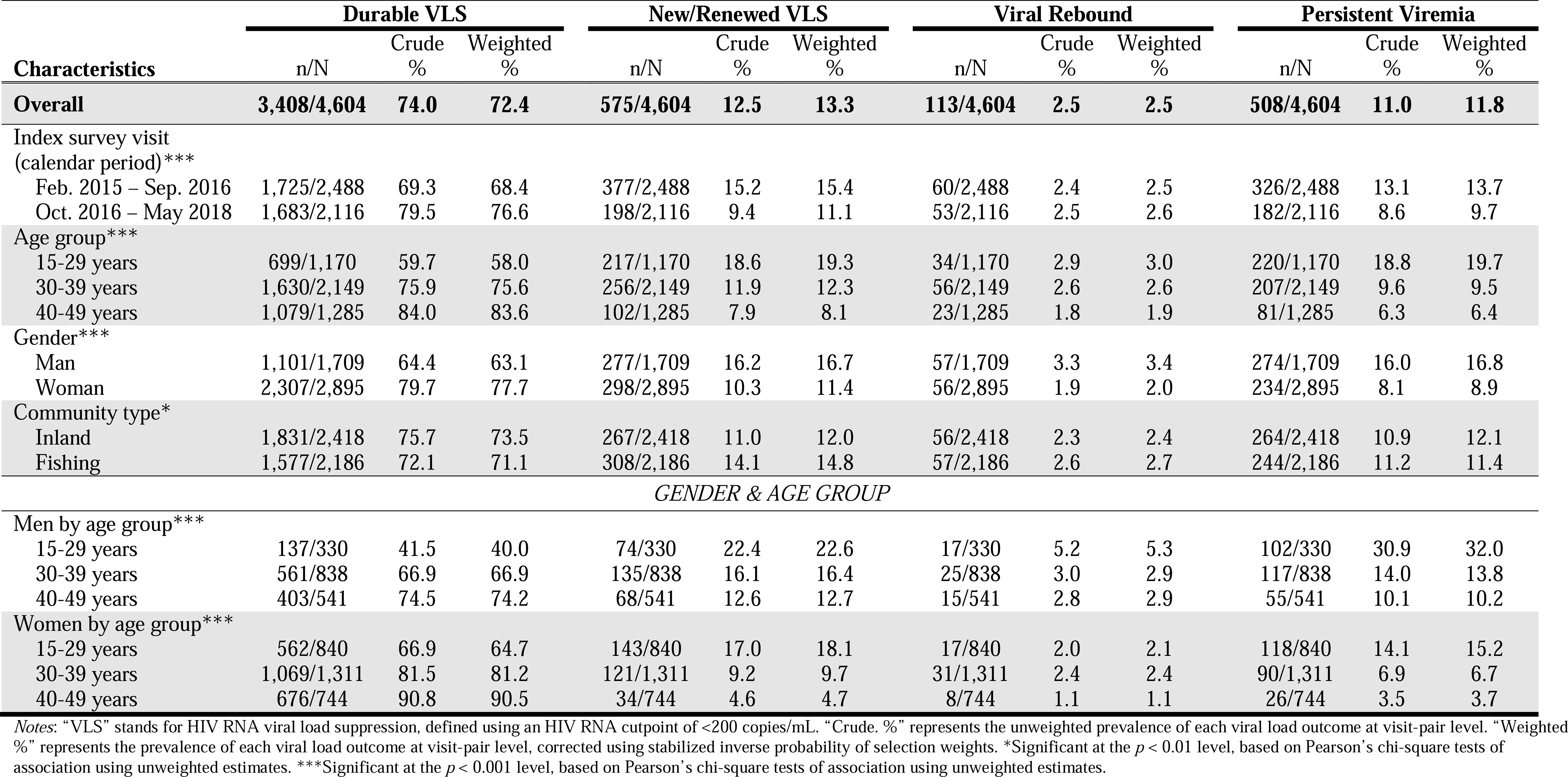
Prevalence of longitudinal virologic outcomes at visit-pair level, by age group, gender, and community type (*N* = 4,604).

| Characteristics | Durable VLS |  |  | New/Renewed VLS |  |  | Viral Rebound |  |  | Persistent Viremia |  |  |
| --- | --- | --- | --- | --- | --- | --- | --- | --- | --- | --- | --- | --- |
|  | n/N | Crude % | Weighted % | n/N | Crude % | Weighted % | n/N | Crude % | Weighted % | n/N | Crude % | Weighted % |
| **Overall** | **3,408/4,604** | **74.0** | **72.4** | **575/4,604** | **12.5** | **13.3** | **113/4,604** | **2.5** | **2.5** | **508/4,604** | **11.0** | **11.8** |
| Index survey visit (calendar period)*** |  |  |  |  |  |  |  |  |  |  |  |  |
| Feb. 2015 – Sep. 2016 | 1,725/2,488 | 69.3 | 68.4 | 377/2,488 | 15.2 | 15.4 | 60/2,488 | 2.4 | 2.5 | 326/2,488 | 13.1 | 13.7 |
| Oct. 2016 – May 2018 | 1,683/2,116 | 79.5 | 76.6 | 198/2,116 | 9.4 | 11.1 | 53/2,116 | 2.5 | 2.6 | 182/2,116 | 8.6 | 9.7 |
| Age group*** |  |  |  |  |  |  |  |  |  |  |  |  |
| 15-29 years | 699/1,170 | 59.7 | 58.0 | 217/1,170 | 18.6 | 19.3 | 34/1,170 | 2.9 | 3.0 | 220/1,170 | 18.8 | 19.7 |
| 30-39 years | 1,630/2,149 | 75.9 | 75.6 | 256/2,149 | 11.9 | 12.3 | 56/2,149 | 2.6 | 2.6 | 207/2,149 | 9.6 | 9.5 |
| 40-49 years | 1,079/1,285 | 84.0 | 83.6 | 102/1,285 | 7.9 | 8.1 | 23/1,285 | 1.8 | 1.9 | 81/1,285 | 6.3 | 6.4 |
| Gender*** |  |  |  |  |  |  |  |  |  |  |  |  |
| Man | 1,101/1,709 | 64.4 | 63.1 | 277/1,709 | 16.2 | 16.7 | 57/1,709 | 3.3 | 3.4 | 274/1,709 | 16.0 | 16.8 |
| Woman | 2,307/2,895 | 79.7 | 77.7 | 298/2,895 | 10.3 | 11.4 | 56/2,895 | 1.9 | 2.0 | 234/2,895 | 8.1 | 8.9 |
| Community type* |  |  |  |  |  |  |  |  |  |  |  |  |
| Inland | 1,831/2,418 | 75.7 | 73.5 | 267/2,418 | 11.0 | 12.0 | 56/2,418 | 2.3 | 2.4 | 264/2,418 | 10.9 | 12.1 |
| Fishing | 1,577/2,186 | 72.1 | 71.1 | 308/2,186 | 14.1 | 14.8 | 57/2,186 | 2.6 | 2.7 | 244/2,186 | 11.2 | 11.4 |
| *GENDER & AGE GROUP* |  |  |  |  |  |  |  |  |  |  |  |  |
| Men by age group*** |  |  |  |  |  |  |  |  |  |  |  |  |
| 15-29 years | 137/330 | 41.5 | 40.0 | 74/330 | 22.4 | 22.6 | 17/330 | 5.2 | 5.3 | 102/330 | 30.9 | 32.0 |
| 30-39 years | 561/838 | 66.9 | 66.9 | 135/838 | 16.1 | 16.4 | 25/838 | 3.0 | 2.9 | 117/838 | 14.0 | 13.8 |
| 40-49 years | 403/541 | 74.5 | 74.2 | 68/541 | 12.6 | 12.7 | 15/541 | 2.8 | 2.9 | 55/541 | 10.1 | 10.2 |
| Women by age group*** |  |  |  |  |  |  |  |  |  |  |  |  |
| 15-29 years | 562/840 | 66.9 | 64.7 | 143/840 | 17.0 | 18.1 | 17/840 | 2.0 | 2.1 | 118/840 | 14.1 | 15.2 |
| 30-39 years | 1,069/1,311 | 81.5 | 81.2 | 121/1,311 | 9.2 | 9.7 | 31/1,311 | 2.4 | 2.4 | 90/1,311 | 6.9 | 6.7 |
| 40-49 years | 676/744 | 90.8 | 90.5 | 34/744 | 4.6 | 4.7 | 8/744 | 1.1 | 1.1 | 26/744 | 3.5 | 3.7 |

Notes: “VLS” stands for HIV RNA viral load suppression, defined using an HIV RNA cutpoint of <200 copies/mL. “Crude. %” represents the unweighted prevalence of each viral load outcome at visit-pair level. “Weighted %” represents the prevalence of each viral load outcome at visit-pair level, corrected using stabilized inverse probability of selection weights. \*Significant at the $p < 0.01$ level, based on Pearson’s chi-square tests of association using unweighted estimates. \*\*\*Significant at the $p < 0.001$ level, based on Pearson’s chi-square tests of association using unweighted estimates.

Adults aged 40-49 years (83.6%) and women (77.7%) were significantly more likely to exhibit durable VLS (*p*<0.001) compared to young persons aged 15-29 years (58.0%) and men (58.0%), respectively. The prevalence of durable VLS was comparable in inland (73.5%) and fishing (71.1%) communities. Of note, young men (41.5%) and women (66.9%) exhibited significantly (*p*<0.001) lower rates of durable VLS relative to older men and women (74.2% and 90.5%), respectively. The prevalence of persistent viremia was twice as high in young men (32.0%) relative to all other age groups, irrespective of gender.

Among participants with three visits (*n* = 1,524), a majority maintained VLS throughout (70.9%) or achieved VLS during (18.6%) the analysis period (**Figure 1**). Few participants exhibited persistent viremia (5.9%), viral rebound (2.6%), or intermittent viremia (1.9%).

### Dynamics of persistent high- and low-level viremia

**Figure 2** shows conditional probabilities of high- and low-level viremia at follow-up given viral load outcomes at the index visit. Among visit-pairs with viremia at the index visit, nearly half (46.9%, 508/1,083) maintained viremia through follow-up. A substantially greater fraction of this viremia was high-level (91.3%, 464/508) as opposed to low-level viremia.

Visit-pairs with high-level viremia at their index visits (*n* = 960) were slightly more likely to achieve VLS (53.0%) than maintain high-level viremia (44.7%) at follow-up, with few (2.3%) progressing to low-level viremia. By comparison, visit-pairs with suppression at the index visit (*n* = 3,521) were substantially more likely to maintain VLS (96.8%) than experience rebounding low- (1.1%) or high-level (2.1%) viremia at follow-up. Likewise, over half of visit-pairs with low-level viremia at their index visits (*n* = 123) achieved VLS at follow-up (53.6%), but substantial fractions maintained low-level viremia (17.9%) or rebounded to high-level viremia (28.5%) at follow-up. Conditional probabilities of high- and low-level viremia were also comparable when estimated separately by calendar period (**Figure S2**) and community type (**Figure S3**), respectively.

Among visit-pairs with persistent high-level viremia (*n* = 429), 20.8% were in persons self-reporting current ART use at their index and follow-up visits (≥12 calendar months apart) (**Table S5**). Among persons self-reporting ART use for ≥12 months, persistent high-level viremia was significantly elevated in younger persons (median age: 33 vs. 36 years), in-migrants (4.7% vs. 2.5%, *p*=0.019), and individuals with ≥2 past-year sexual partners (4.0% vs. 2.4%, *p*=0.021) relative to durable VLS (**Table S6**).

### Temporal and geographic patterns of persistent high-level viremia

Significant increases (from 68.4% pre-UTT scale-up to 76.6% post-UTT scale-up) in the prevalence of durable VLS were accompanied by reductions in persistent high-level viremia (from 13.7% to 9.7%, adjusted risk ratio [adjRR] = 0.75, 95% confidence interval [95%CI] 0.66-0.86) (**Table 1**, **Figure 3**). Significant reductions in persistent high-level viremia over calendar period were also observed across population strata (men: adjRR=0.70, 95%CI 0.59-0.83; women: adjRR=0.81, 95%CI 0.67-0.98) and community types (inland: adjRR=0.83, 95%CI 0.70-1.00; fishing: adjRR=0.69, 95%CI 0.57-0.84) (**Figure 3**).

Nevertheless, changes in the prevalence of persistent high-level viremia over time were not uniform, with some geographic areas exhibiting greater declines than others. Within regions, for example, the prevalence of persistent high-level viremia declined in all but one region (**Figure 4A-4B**), and the magnitude of these declines varied across regions (**Table S7**). Observed declines in persistent viremia were primarily driven by increased VLS in men over time, but men exhibited substantially higher fractions of persistent viremia than women (**Figure 4A-4B**). Community-level analyses revealed heterogeneities in the magnitudes of decline in persistent viremia over calendar time (**Figure 4C**, **Table S8**). Across communities, however, observed declines in persistent high-level viremia were generally not statistically significant (**Table S8**), and community-level variations in the magnitude of these declines were unexplained by differences in community type (**Figure 4C**).

### Individual-level predictors of persistent high-level viremia

**Table 2** presents unweighted and weighted associations for persistent high-level viremia, relative to sustained or new/renewed VLS. Overall, men (adjRR=2.40, 95%CI 1.87-3.07) exhibited significantly higher risk of persistent viremia relative to women. Secondary education (adjRR=1.51, 95%CI 1.17-1.95), inconsistent condom use with casual partners (adjRR=1.38, 95%CI 1.10-1.74), and hazardous alcohol use (adjRR=1.09, 95%CI 1.03-1.16) were also significantly associated with persistent high-level viremia. In-migrants were also substantially, but not significantly, more likely than long-term residents to exhibit persistent high-level viremia (adjRR=1.51, 95%CI 0.99-1.44).

**Table 2.** Unweighted and weighted risk of persistent high-level viremia (≥1,000 copies/mL) relative to sustained or new/renewed low-level viremia or suppression (*N* = 3,028).

| Characteristics | Unweighted |  | Weighted |  |
| --- | --- | --- | --- | --- |
|  | RR (95% CI) | adjRR (95% CI) | RR (95% CI) | adjRR (95% CI) |
| **Demographics** |  |  |  |  |
| Age group |  |  |  |  |
| 15-29 years | **3.08 (2.34–4.05)** | **2.81 (2.12–3.73)** | **3.12 (2.34–4.15)** | **2.96 (2.21–3.96)** |
| 30-39 years | **1.62 (1.25–2.11)** | **1.57 (1.21–2.03)** | **1.65 (1.26–2.17)** | **1.63 (1.24–2.15)** |
| 40-49 years | *Ref.* | *Ref.* | *Ref.* | *Ref.* |
| Gender |  |  |  |  |
| Man | **2.26 (1.84–2.77)** | **2.57 (2.02–3.27)** | **2.04 (1.63–2.53)** | **2.40 (1.87–3.07)** |
| Woman | *Ref.* | *Ref.* | *Ref.* | *Ref.* |
| Currently marital status |  |  |  |  |
| Never married | **1.82 (1.37–2.42)** | **1.34 (1.02–1.79)** | **1.78 (1.34–2.37)** | 1.28 (0.96–1.70) |
| Currently married | *Ref.* | *Ref.* | *Ref.* | *Ref.* |
| Previously married | 0.94 (0.77–1.15) | 1.02 (0.84–1.25) | 0.97 (0.78–1.22) | 1.04 (0.84–1.29) |
| Completed education |  |  |  |  |
| No formal education | 0.78 (0.53–1.15) | 0.91 (0.62–1.32) | 0.84 (0.57–1.25) | 1.02 (0.70–1.50) |
| Primary | *Ref.* | *Ref.* | *Ref.* | *Ref.* |
| Secondary | **1.31 (1.02–1.68)** | **1.36 (1.07–1.74)** | **1.43 (1.09–1.88)** | **1.51 (1.17–1.95)** |
| Technical/University | 0.74 (0.34–1.62) | 1.02 (0.48–2.18) | 0.70 (0.31–1.55) | 1.00 (0.45–2.23) |
| Primary occupation |  |  |  |  |
| Agriculture or housework | *Ref.* | *Ref.* | *Ref.* | *Ref.* |
| Trading or shopkeeping | 1.10 (0.88–1.38) | 0.99 (0.79–1.24) | 1.16 (0.84–1.33) | 0.96 (0.77–1.21) |
| Bar/sex work or waitressing | 0.75 (0.52–1.08) | 0.80 (0.55–1.18) | 0.69 (0.47–1.01) | 0.75 (0.49–1.14) |
| Fishing-related occupation | **1.83 (1.44–2.32)** | 0.98 (0.75–1.29) | **1.58 (1.22–2.04)** | 0.89 (0.67–1.18) |
| Other | 1.23 (0.98–1.55) | 0.96 (0.48–1.21) | 1.14 (0.88–1.47) | 0.93 (0.73–1.19) |
| Household wealth (quartile) |  |  |  |  |
| Lowest | **1.43 (1.09–1.89)** | 1.27 (0.96–1.69) | **1.36 (1.00–1.86)** | 1.24 (0.91–1.70) |
| Low-middle | **1.36 (1.02–1.81)** | 1.14 (0.85–1.52) | 1.27 (0.92–1.76) | 1.09 (0.79–1.51) |
| High-middle | **1.39 (1.07–1.83)** | 1.29 (0.99–1.68) | 1.33 (0.96–1.84) | 1.22 (0.90–1.64) |
| Highest | *Ref.* | *Ref.* | *Ref.* | *Ref.* |
| Migration |  |  |  |  |
| Long-term resident | *Ref.* | *Ref.* | *Ref.* | *Ref.* |
| In-migrant | **1.48 (1.23–1.79)** | **1.26 (1.04–1.52)** | **1.51 (1.24–1.82)** | 1.19 (0.99–1.44) |
| **Behavioral** |  |  |  |  |
| Number of sexual partners |  |  |  |  |
| 0–1 | *Ref.* | *Ref.* | *Ref.* | *Ref.* |
| $\geq 2$ | **1.47 (1.23–1.75)** | 0.82 (0.67–1.01) | **1.42 (1.17–1.72)** | 0.83 (0.67–1.03) |
| Condom use |  |  |  |  |
| No partners or permanent partners only | *Ref.* | *Ref.* | *Ref.* | *Ref.* |
| Consistent use with casual partners | **1.44 (1.17–1.79)** | 1.19 (0.95–1.50) | **1.51 (1.19–1.91)** | 1.26 (0.99–1.61) |
| Inconsistent use with casual partners | **1.62 (1.34–1.98)** | **1.43 (1.16–1.77)** | **1.60 (1.29–1.99)** | **1.38 (1.10–1.74)** |
| Hazardous alcohol use | **1.15 (1.09–1.21)** | **1.07 (1.01–1.13)** | **1.16 (1.09–1.24)** | **1.09 (1.03–1.16)** |
| Intimate partner violence |  |  |  |  |
| None | *Ref.* | *Ref.* | *Ref.* | *Ref.* |
| Any | **1.21 (1.03–1.44)** | 1.03 (0.87–1.22) | **1.23 (1.04–1.46)** | 1.05 (0.88–1.24) |

Notes: Risk ratios (RR) and 95% confidence intervals (95%CI) were estimated using Poisson regression with generalized estimating equations, exchangeable covariance matrices, and robust standard errors. Multivariable models were adjusted for index survey interval and all covariates displayed in the columns presenting adjusted results. Sustained or new/renewed low-level viremia or suppression was defined as $<1,000$ copies/mL across visits or at follow-up only. **Bolded** values represent risk ratios of persistent HIV viremia that were significantly different from the null value of 1 at the $p < 0.05$ level or below. Behavioral factors were measured in the past year.

Weighted multivariable models stratified by gender and community type, respectively, yielded similar findings. However, persistent viremia was associated with inconsistent condom use with casual partners (adjRR=1.38, 95%CI 1.05-1.80) and hazardous alcohol use (adjRR=1.08, 95%CI 1.01-1.16) in men but not women (**Table S11**). Likewise, household poverty (adjRR=1.59, 95%CI: 1.09-2.33) and hazardous alcohol use (adjRR=1.17, 95%CI 1.08-1.27) were associated with higher risk of persistent viremia in inland communities only (**Table S12**).

## Discussion

This population-based study in 36 moderate-prevalence inland communities and four high-prevalence fishing communities in southern Uganda demonstrated significant increases in durable HIV VLS coinciding with reductions in persistent viremia during UTT scale-up. Similar declines in persistent viremia were observed by gender and age as well as across community types, suggesting UTT rollout conferred benefits across population strata and geographic settings. Despite the large fraction of persons exhibiting durable VLS or new/renewed suppression, persistent high-level viremia remained prevalent in the presence of UTT. Persistent high-level viremia was also most frequently observed in specific groups – namely youth, men, in-migrants, and individuals with higher-risk behaviors (i.e., inconsistent condom use with casual partners, hazardous alcohol use) – who experience substantial barriers along the HIV care continuum, from diagnosis to retention in care, throughout sub-Saharan Africa.^27,28^ While UTT may accelerate progress towards HIV epidemic control goals, our findings reveal substantive disparities in longitudinal virologic outcomes amidst UTT scale-up. These disparities are most pronounced in groups with greatest potential for onward HIV transmission,^29–32^

A noteworthy fraction (∼20%) of persistent high-level viremia was observed among individuals self-reporting ART use for ≥12 months, which could reflect unaddressed virologic failure and/or suboptimal ART adherence in this population. Factors associated with persistent viremia among persons on ART reported elsewhere include HIV drug resistance, delayed switching to second- or third-line ART regimens, viral load monitoring gaps, and poor care quality.^33–35^ Point-of-care viral load monitoring may reduce delays in detecting and responding to virologic failure among persons on ART exhibiting viremia for >12 months.^36^ Furthermore, persistently viremic men were significantly more likely than women to have never initiated ART, which could reflect gendered patterns in anticipated/enacted HIV stigma, masculine norms surrounding care-seeking, and perceptions that health facilities are predominantly female-oriented spaces.^37,38^ Collectively, these forces may dissuade men from initiating and continuing HIV care.

Despite observed increases in durable VLS coinciding with UTT scale-up, declines in persistent high-level viremia were not consistent across communities, revealing geographic heterogeneities in the impact of UTT rollout. Although some of these differences could be explained by demographics, they were unexplained by community type alone, as declines varied even among communities of the same type. Unobserved factors beyond community type, including facility-level attributes and HIV-1 drug resistance, could be driving these observed geographic heterogeneities in persistent viremia over time.^39,40^ Future research should continue investigating these factors to identify areas for potential health system interventions with potential to accelerate the impact of UTT on population-level VLS and HIV epidemic dynamics.

Persistent high-level viremia in this study was significantly more common among youth, men, in-migrants, and individuals with higher-risk behavioral profiles (i.e., inconsistent condom use, hazardous alcohol use). These populations have been shown to shoulder disproportionate burdens of viremia, both in this study setting and elsewhere.^12,23,41,42^ Of concern, over a third young men exhibited persistent high-level viremia—twice the rate observed in young women. Given that young men in this setting are highly mobile and report substantial barriers to HIV care linkage and retention (e.g., obtaining facility transfer forms),^43–45^ expanding eligibility for lower-intensity differentiated service delivery models (i.e., multi-month ART dispensing, fast-track drug refills) to some persistently viremic persons and enhancing efficiencies in the facility transfer process are potential solutions to improving virologic outcomes in these groups. Enhanced linkage-to-care efforts and novel therapeutics like long-acting ART may also optimize clinical outcomes in these populations.

This is the first study to characterize population-level patterns and predictors of durable VLS and persistent high- and low-level viremia after UTT in both hyperendemic and moderate-prevalence communities in sub-Saharan Africa. Nevertheless, study findings are subject to several limitations. First, loss to follow-up in the study population was substantial, and non-participation was associated with in- and out-migration, which are known correlates of viremia. Estimates of persistent viremia and viral rebound in the analytic cohort, therefore, likely underestimate the true prevalence of viremia in the source population. Although inverse probability weights helped address potential attrition biases induced by excluding individuals with only one study visit, this procedure attempts to balance the analytic cohort and the excluded population on measured *and* unobserved factors, which requires assumptions of exchangeability that are difficult to assess.^46,47^ Second, ART use and other behavioral factors were self-reported and, thus, are susceptible to recall and acquiescence biases. Third, the present study did not include measures of ART adherence, ART use duration, HIV treatment interruptions, ART regimens, or HIV drug resistance—all of which are known predictors of longitudinal HIV viremia. Fourth, the interval between study visits was prolonged (≥12 calendar months apart), and VLS was ascertained only at the start and end of each interval. We, therefore, lacked more repeated, frequent measurements of viremia throughout visit intervals. Lastly, our study focused on demographic, behavioral, and geographic predictors of persistent viremia and did not address the contribution of health systems factors to longitudinal virologic outcomes.

## Conclusions

This study in 40 continuously surveilled communities with heterogeneous HIV burdens in south-central Uganda documented significant increases in durable VLS during UTT scale-up. Nevertheless, noteworthy individual and geographic disparities in persistent high-level viremia coincided with population-level gains in durable VLS, which clustered disproportionately in older adults and women. Individuals with viremia were equally as likely to achieve VLS or maintain viremia. Youth, men, in-migrants, and individuals reporting higher-risk behaviors (i.e., inconsistent condom use with casual partners, hazardous alcohol use) exhibited significantly elevated risk of persistent high-level viremia during UTT scale-up, suggesting universal ART provision alone does not close gaps in virologic outcomes in populations at greatest risk for onward HIV transmission. To reach ambitious treatment targets and achieve HIV epidemic control, complementary treatment optimization interventions (e.g., enhanced linkage-to-care, expedited viral load testing, long-acting ART introduction, sustained geographic prioritization, streamlining facility transfer processes) must be directed at populations who remain persistently viremic despite UTT.

## Competing interests

The authors have no conflicts of interest to disclose.

## Supporting information

Electronic Supplementary Material

## Data Availability

The data used in the present study can be requested for replication purposes or alternative secondary analyses from the senior author, Dr. M. Kate Grabowski.

## Acknowledgements

We are grateful to all RCCS participants for their time and contributions to the present study. We also acknowledge the dedication and efforts of all data collection, laboratory, service linkage, and data management staff. Lastly, we thank Maria Wawer and Ron Gray for their review of study objectives and their input into the analysis plan and results interpretation.

## Funding

This study was supported by the National Institute of Allergy and Infectious Diseases (U01AI100031, U01AI075115, R01AI110324, R01AI128779, R01AI123002, R01AI143333, R01AI55080, T32AI102623); the National Institute of Mental Health (R01MH105313); the National Cancer Institute (75N91019D00024); and the U.S. President’s Emergency Plan for AIDS Relief through the Centers for Disease Control and Prevention under the terms of (NU2GGH000817). JGR and KBR were supported by the National Institute of Mental Health (F31MH126796, K01MH129226). EUP was supported by the National Institute on Drug Abuse (F31DA054849).

## Disclaimer

The contents of this manuscript are solely the responsibility of the authors and do not necessarily reflect the views or policies of the funding agencies, nor do mentions of trade names, commercial products, or organizations imply endorsement by the U.S. Government.

## Supporting information

Supporting Information File 1: Electronic Supplementary Material

DOCX. File contains supplemental tables and figures for the present manuscript.

## List of abbreviations

ART: Antiretroviral therapy
CI: Confidence interval
mL: Milliliter
QIC: Quasilikelihood under the independence model criteria
RCCS: Rakai Community Cohort Study
RR: Risk ratio
UTT: Universal Test and Treat
VLS: Viral load suppression

---

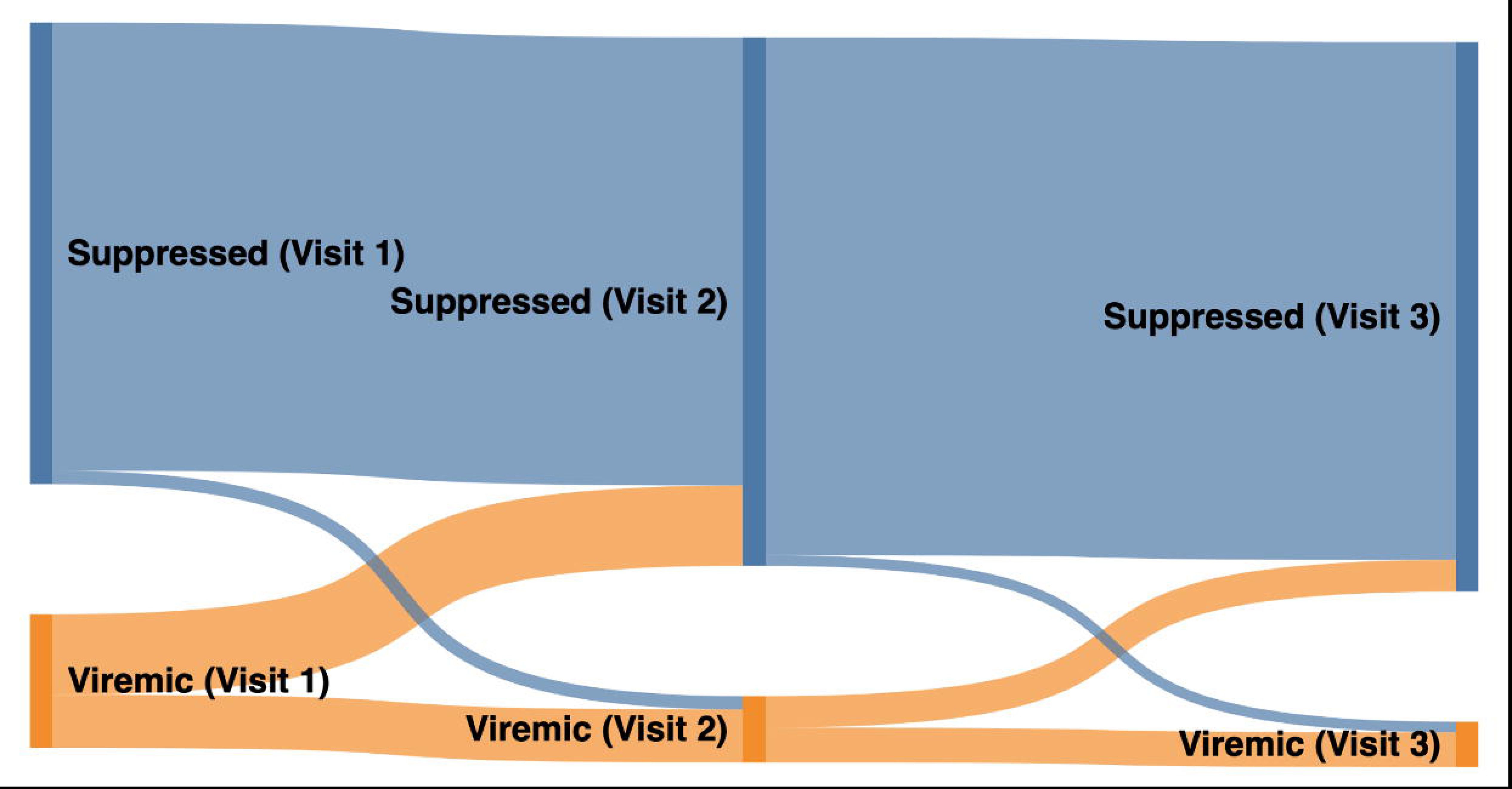

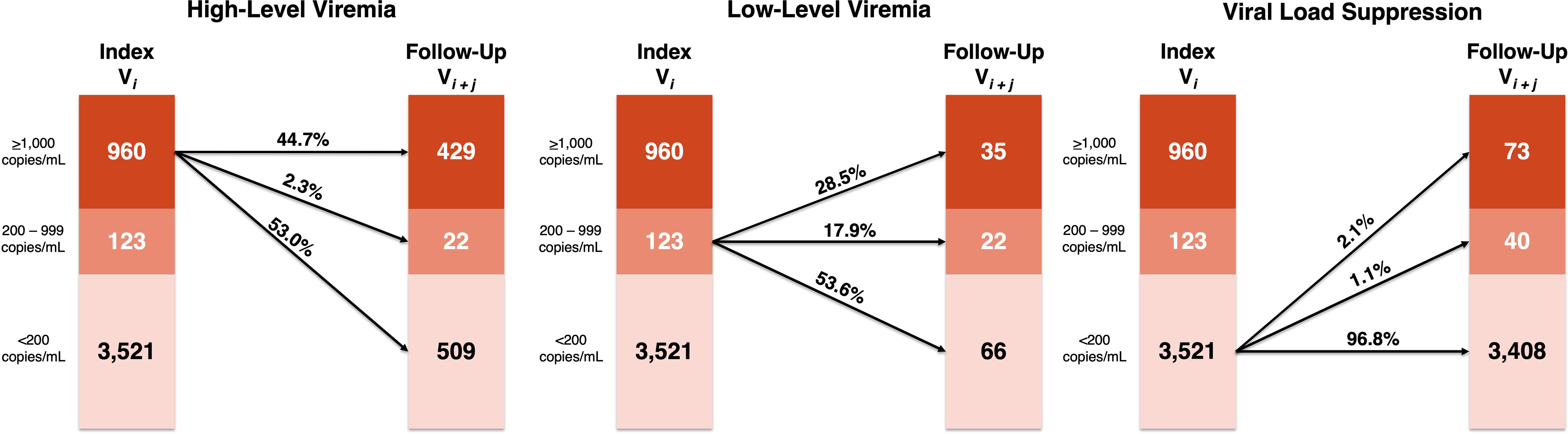

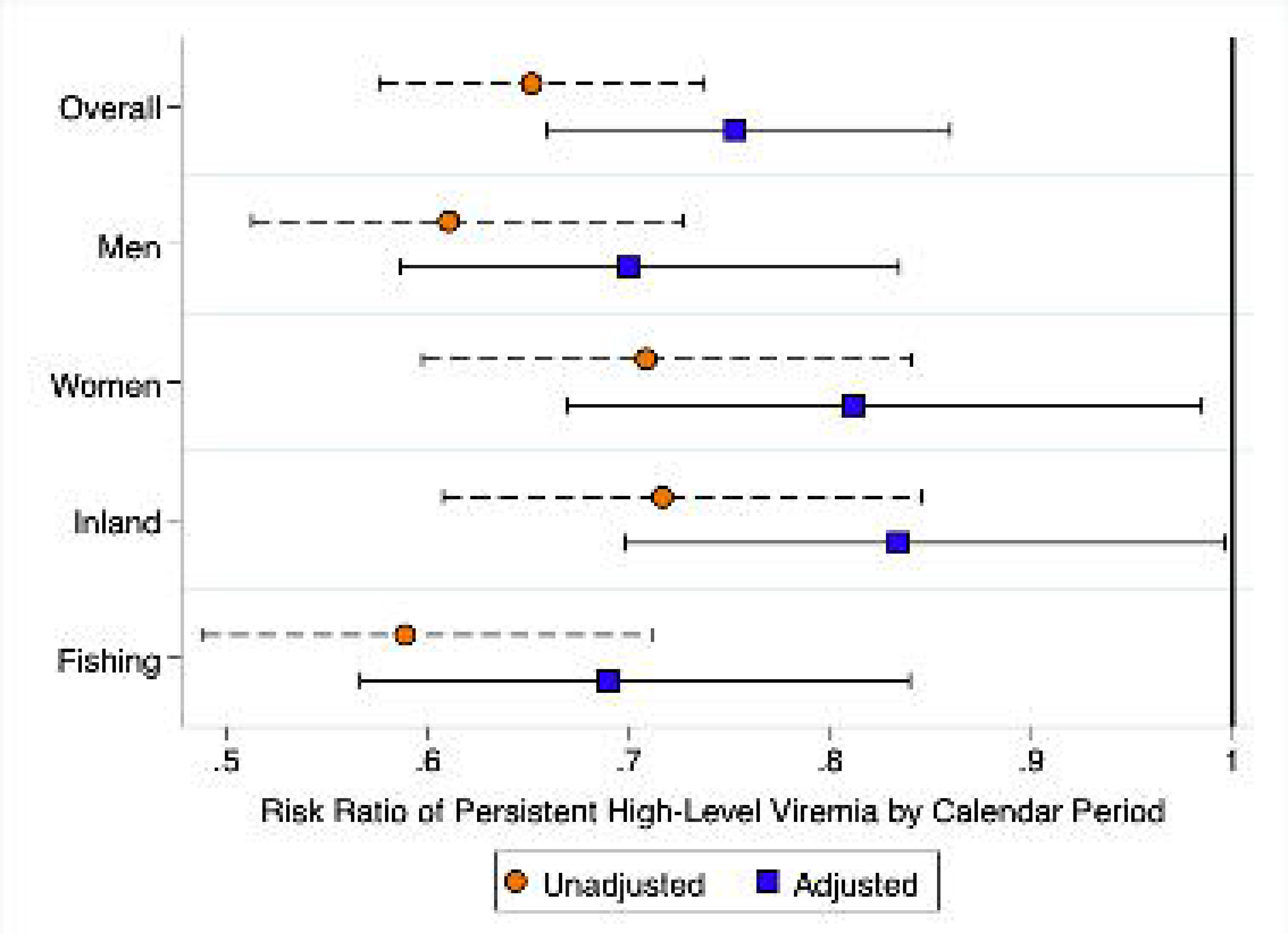

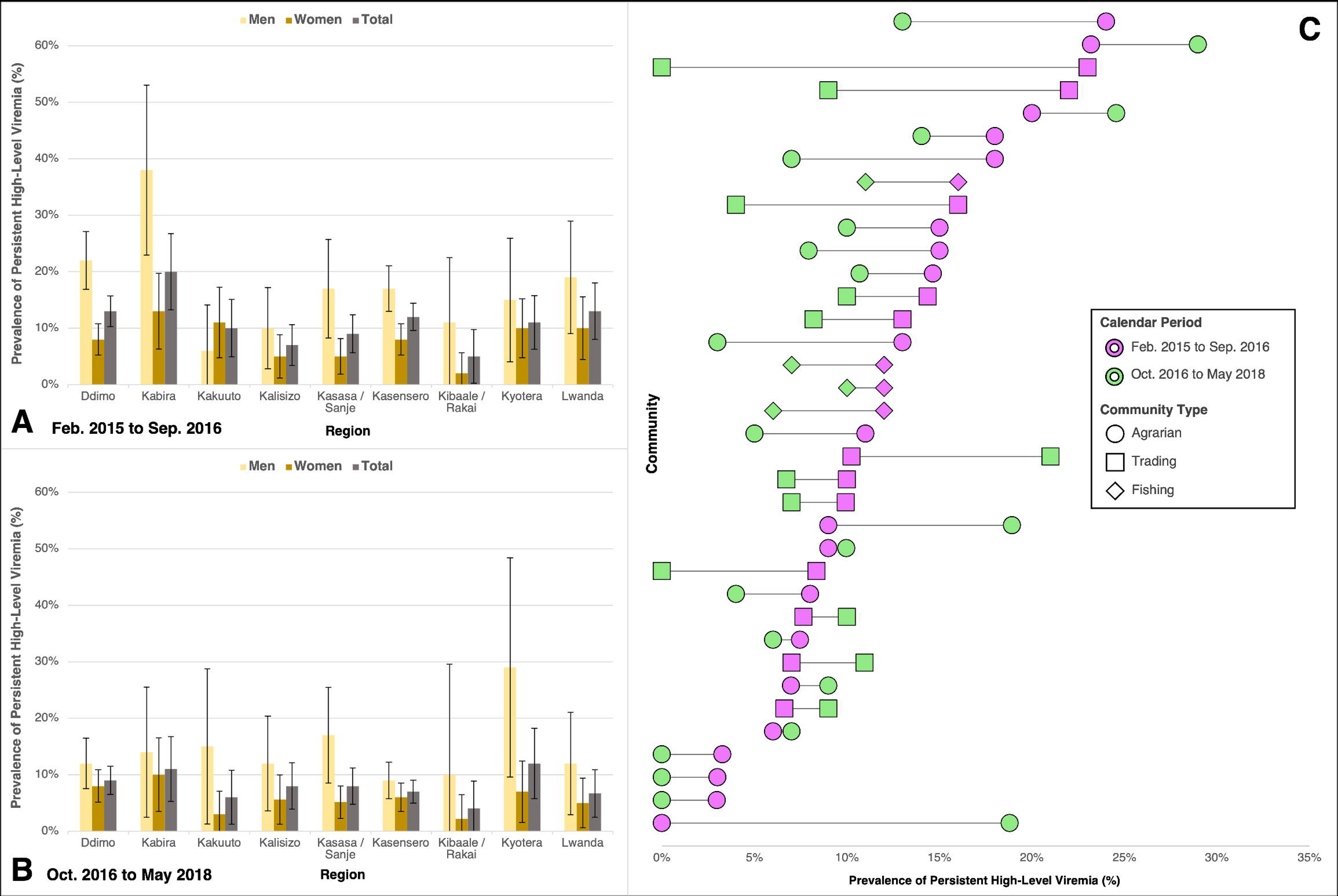

## Notes

### Competing Interest Statement

The authors have declared no competing interest.

### Author Declarations

The Uganda Virus Research Institute Research and Ethics Committee and the Johns Hopkins University School of Medicine Institutional Review Board reviewed and approved the study protocol. Adults (>18 years) and emancipated minors provided written informed consent prior to study procedures. Written assent and parental consent were obtained for unemancipated minors aged 15-17 years.

