## Supplementary material for "Temporal dynamics and drivers of durable HIV viral load suppression and persistent high- and low-level viremia during Universal Test and Treat scale-up in Uganda: a population-based study": Electronic Supplementary Material

**Table S1.** Measurement and definition of independent variables from the Rakai Community Cohort Study (RCCS) included in analysis.

| **Variable** | **Type** | **Definition** |
| --- | --- | --- |
| Educational attainment | Categorical | Level of schooling, complete or incomplete, from primary 1–7 (*Primary*); secondary 1–6 (*Secondary*); or technical/university, primary professional, O’level professional, primary/O’level apprenticeship, or A’level apprenticeship (*Technical/University*). Participants who reported never attending school were classified as having *No Formal Education.* |
| Primary occupation | Categorical | Activities or work that keep participants busy on average day (for money or not), collapsed into the following categories: agriculture for home use/bartering/selling, housework in own home, housekeeping in someone else’s home, or home brewing (*Agriculture or Housework*); shopkeeping, trading/vending, or hairdresser/salon owner (*Trading or Shopkeeping*); bar worker/owner, waitress/waiter/restaurant owner, or sex worker (*Bar Work, Waitressing, or Sex Work*); fishing (*Fishing-Related Occupation*); or government/clerical/teaching, student, military/police, medical worker, casual laborer, construction, mechanic, transportation (trucker/boda boda), sports betting, or unemployed (*Other*). |
| Religion | Categorical | Self-reported religious identity, grouped by the following: Catholic, Protestant, Church of Uganda, Saved/Pentecostal (*Catholic/Christian*); *Muslim*; or *Other/None*. |
| Household wealth | Categorical | Enumerated from an aggregation of 9 household possessions and dwelling characteristics (yes or no) and partitioned into quartiles (groups of four) standardized at each survey round, as described by Santelli *et al*. (2021).^1^ |
| Migration | Categorical | Self-reported migration into an RCCS community from outside the study since the prior round (~18 calendar months) (*In-Migrants*) compared to individuals who did not migrate into the study area in the prior round (*Long-Term Residents*). |
| Condom use | Categorical | Sometimes or never (*Inconsistent*) versus always (*Consistent*) using condoms in the past year with any non-marital/casual partner (i.e., visitor, stranger, workmates/colleague, boss/work supervisor, employee, or sugar daddy/mummy). Those reporting no partners or marital/permanent partners only (i.e., current/former spouses or long-term partners) in the past year served as the referent group. |
| Transactional sex | Categorical | Giving and/or receiving money, gifts, or favors in exchange for sex in the past year with any partner (yes or no). |
| Hazardous alcohol use | Count | Aggregated number of reported experiences in the past year following alcohol consumption, adapted from Miller *et al* (2021):^2^ (1) unsteady gait, (2) fell over, (3) got angry, (4) got violent/into a fight, (5) difficulty speaking, (6) forgot events while drinking, (7) shaking hands the next morning, (8) felt ashamed. Participants reporting no alcohol consumption in the past year received scores of 0. |
| Any alcohol use consequences | Categorical | Reported any of the following consequences following alcohol use in the past year (any or none/no alcohol consumption), adapted from Miller *et al* (2021):^2^ (1) unsteady gait, (2) fell over, (3) got angry, (4) got violent/into a fight, (5) difficulty speaking, (6) forgot events while drinking, (7) shaking hands the next morning, (8) felt ashamed. |
| Illicit drug use | Categorical | Used any of the following illicit substances in the past year (yes or no): marijuana, amphetamines, aero fuels (“glue”), amayirungi (“khat”), heroin, or kuber. |
| Intimate partner violence* | Categorical | Reported perpetrating or experiencing any of the following with any partner in the past year (yes or no): (1) verbally abused/shouted; (2) pushed, slapped, or held down; (3) punched with something that could injure; (4) kicked/dragged; (5) threatened with a weapon (knife, gun, fire, rope); (6) used threats to force someone to have sex; (7) physically forced to have sex; (8) forced to perform sexual acts against will. Items were derived from the Revised Conflicts Tactics Scale.^3^ |

**Variable first introduced during the Round 18 survey interval (October 2016 – May 2018).*

**Figure S1.** Flow chart of inclusion into the analytic cohort from the Rakai Community Cohort Study (RCCS).

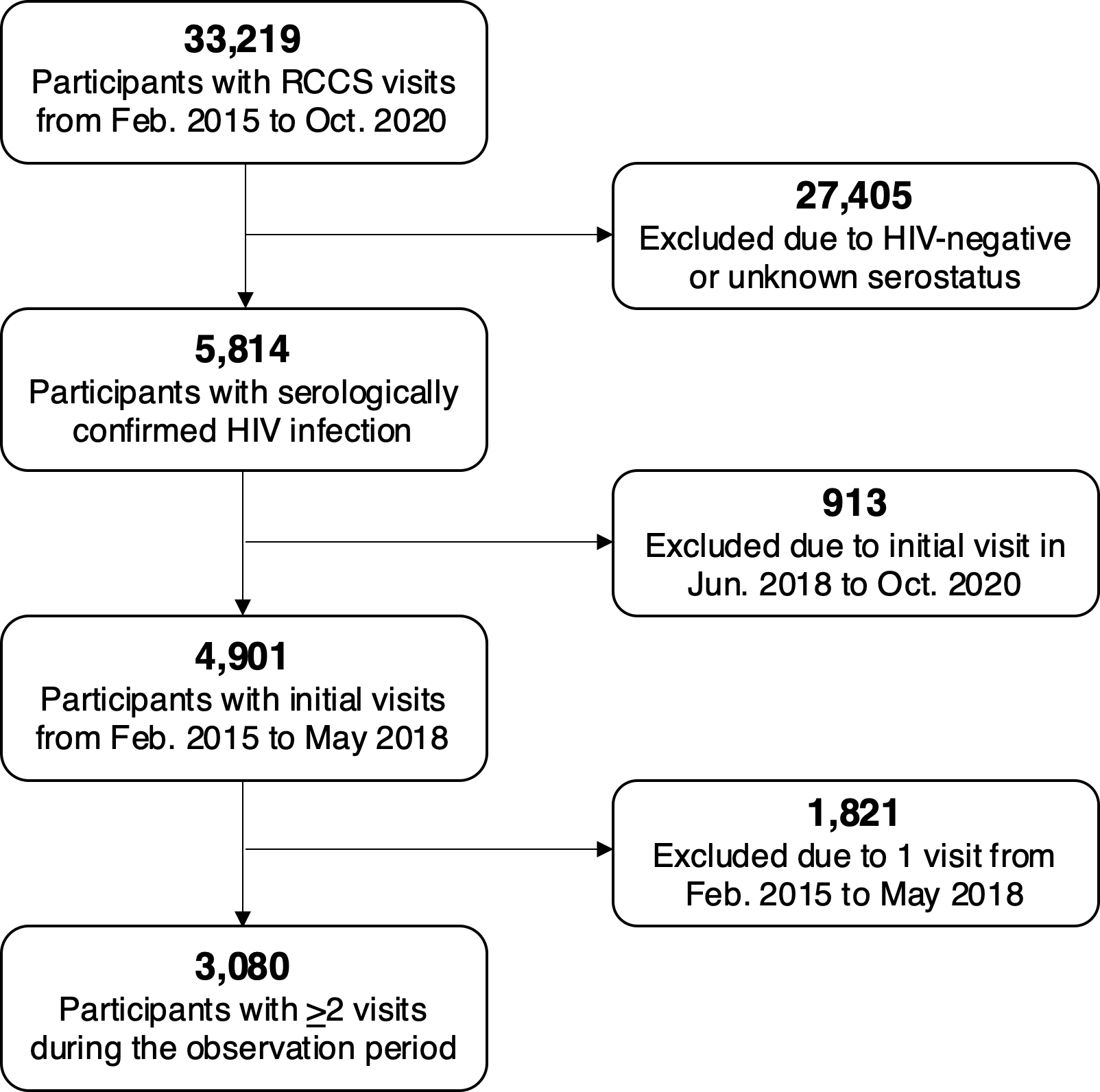

**Table S2**. Descriptive sample statistics at the index visit comparing participants with ≥2 visits (analytic cohort) to those with 1 visit (lost to follow-up) during the observation period—2015 to 2020.

| **Characteristics (*n*, %)** | **Total**  *N* = 4,901 |  | **Lost to Follow-Up**  *n* = 1,821 (37.2%) | **Analytic Cohort**  *n* = 3,080 (62.8%) | **χ²**  ***p*-value*** |
| --- | --- | --- | --- | --- | --- |
| ***Demographics*** |  |  |  |  |  |
| Index survey visit (calendar period) |  |  |  |  | **<0.001** |
| Feb. 2015 – Sep. 2016 | 3,606 (73.6) |  | 1,118 (61.4) | 2,488 (80.8) |  |
| Oct. 2016 – May 2018 | 1,295 (26.4) |  | 703 (38.6) | 592 (19.2) |  |
| Age, in years (*median*, *IQR*)† | 33 (28–39) |  | 31 (26–38) | 34 (28–39) | **<0.001** |
| Age group |  |  |  |  | **<0.001** |
| 15-29 years | 1,688 (34.4) |  | 780 (42.8) | 908 (29.5) |  |
| 30-39 years | 2,102 (42.9) |  | 681 (37.4) | 1,421 (46.1) |  |
| 40-49 years | 1,111 (22.7) |  | 360 (19.8) | 751 (24.4) |  |
| Gender |  |  |  |  | 0.226 |
| Man | 1,835 (37.4) |  | 662 (36.3) | 1,173 (38.1) |  |
| Woman | 3,066 (62.6) |  | 1,159 (63.7) | 1,907 (61.9) |  |
| Currently marital status |  |  |  |  | 0.131 |
| Never married | 374 (7.6) |  | 153 (8.4) | 221 (7.2) |  |
| Currently married | 2,855 (58.3) |  | 1,032 (56.7) | 1,823 (59.2) |  |
| Previously married | 1,672 (34.1) |  | 636 (34.9) | 1,036 (33.6) |  |
| Educational attainment |  |  |  |  | 0.990 |
| No formal education | 412 (8.4) |  | 152 (8.4) | 260 (8.5) |  |
| Primary | 3,581 (73.1) |  | 1,329 (73.0) | 2,252 (73.1) |  |
| Secondary | 787 (16.0) |  | 296 (16.2) | 491 (15.9) |  |
| Technical/University | 121 (2.5) |  | 44 (2.4) | 77 (2.5) |  |
| Primary occupation |  |  |  |  | **<0.001** |
| Agriculture or housework | 1,706 (34.8) |  | 605 (33.2) | 1,101 (35.8) |  |
| Trading or shopkeeping | 1,037 (21.2) |  | 400 (22.0) | 637 (20.7) |  |
| Bar work, waitressing, or sex work | 468 (9.6) |  | 207 (11.4) | 261 (8.5) |  |
| Fishing-related occupation | 768 (15.7) |  | 249 (13.7) | 519 (16.8) |  |
| Other | 922 (18.8) |  | 360 (19.8) | 562 (18.2) |  |
| Religion |  |  |  |  | 0.398 |
| Catholic/Christian | 4,291 (87.6) |  | 1,587 (87.1) | 2,704 (87.8) |  |
| Muslim | 572 (11.7) |  | 216 (11.9) | 356 (11.6) |  |
| Other or none | 38 (0.8) |  | 18 (1.0) | 20 (0.6) |  |
| Household wealth (quartile) |  |  |  |  | **<0.001** |
| Lowest | 2,015 (41.1) |  | 684 (37.6) | 1,331 (43.2) |  |
| Low-middle | 1,046 (21.3) |  | 392 (21.5) | 654 (21.2) |  |
| High-middle | 1,097 (22.4) |  | 427 (23.4) | 670 (21.8) |  |
| Highest | 728 (14.9) |  | 309 (17.0) | 419 (13.6) |  |
| *Missing* | *15 (0.3)* |  | *9 (0.5)* | *6 (0.2)* |  |
| Migration |  |  |  |  | **<0.001** |
| Long-term resident | 3,510 (71.6) |  | 1,056 (58.0) | 2,454 (79.7) |  |
| In-migrant | 1,389 (28.3) |  | 763 (41.9) | 626 (20.3) |  |
| *Missing* | *2 (0.1)* |  | *2 (0.1)* | *n/a* |  |
| Community type |  |  |  |  | **<0.001** |
| Agrarian | 1,457 (29.7) |  | 524 (28.8) | 933 (30.3) |  |
| Trading | 1,160 (23.7) |  | 509 (27.9) | 651 (21.1) |  |
| Fishing | 2,284 (46.6) |  | 788 (43.3) | 1,496 (48.6) |  |
| ***Behavioral^§^*** |  |  |  |  |  |
| Number of sexual partners |  |  |  |  | **0.002** |
| 0–1 | 3,359 (68.5) |  | 1,196 (65.7) | 2,163 (70.2) |  |
| ≥2 | 1,542 (31.5) |  | 625 (34.4) | 917 (29.8) |  |
| Condom use |  |  |  |  | 0.067 |
| No partners or permanent partners only | 3,310 (67.5) |  | 1,193 (65.5) | 2,117 (68.8) |  |
| Consistent use with casual partners | 698 (14.2) |  | 275 (15.1) | 423 (13.7) |  |
| Inconsistent use with casual partners | 893 (18.2) |  | 353 (19.4) | 540 (17.5) |  |
| Transactional sex |  |  |  |  | **0.017** |
| No | 2,381 (47.3) |  | 821 (45.1) | 1,497 (48.6) |  |
| Yes | 2,583 (52.7) |  | 1,000 (54.9) | 1,583 (51.4) |  |
| Any alcohol use consequences |  |  |  |  | **0.030** |
| No | 3,655 (74.6) |  | 1,326 (72.8) | 2,329 (75.6) |  |
| Yes | 1,246 (25.4) |  | 495 (27.2) | 751 (24.4) |  |
| Illicit drug use |  |  |  |  | 0.372 |
| No | 4,655 (95.0) |  | 1,723 (94.6) | 2,932 (95.2) |  |
| Yes | 246 (5.0) |  | 98 (5.4) | 148 (4.8) |  |
| ***HIV-related*** |  |  |  |  |  |
| ART use history (self-reported) |  |  |  |  | **<0.001** |
| Never | 1,706 (34.8) |  | 775 (42.6) | 931 (30.2) |  |
| Currently or previously | 3,195 (65.2) |  | 1,046 (57.4) | 2,149 (69.8) |  |
| **HIV RNA viral load, in copies/mL** |  |  |  |  |  |
| Geometric mean viral load  (*95%CI*)† | 7,792  (7,061–8,599) |  | 8,054  (6,941–9,344) | 7,581  (6,645–8,648) | 0.551 |
| <1,000 copies/mL | 3,450 (70.4) |  | 1,160 (63.7) | 2,290 (74.4) | **<0.001** |
| <400 copies/mL | 3,324 (67.8) |  | 1,106 (60.7) | 2,218 (72.0) | **<0.001** |
| <200 copies/mL | 3,268 (66.7) |  | 1,079 (59.3) | 2,189 (71.1) | **<0.001** |
| <50 copies/mL | 3,121 (63.7) |  | 1,011 (55.5) | 2,110 (68.5) | **<0.001** |

**p*-values calculated using Pearson’s chi-square tests of association, unless otherwise specified. † *p*-value calculated using Wilcoxon rank-sum tests comparing

median values and interquartile ranges (IQR). *^§^* Behavioral factors measured in the past year.

**Table S3.** Covariate distributions comparing the unweighted analytic cohort to the inverse probability-weighted pseudo-population.

| **Characteristics (%)** | **Unweighted**  **Analytic Cohort** | **Weighted Pseudo-**  **Population** |
| --- | --- | --- |
| ***Demographics*** |  |  |
| Index survey visit (calendar period) |  |  |
| Feb. 2015 – Sep. 2016 | 80.8 | 73.5 |
| Oct. 2016 – May 2018 | 19.2 | 26.5 |
| Age, in years (*mean*) | 33.8 | 33.1 |
| Age group |  |  |
| 15-29 years | 29.5 | 34.7 |
| 30-39 years | 46.1 | 42.8 |
| 40-49 years | 24.4 | 22.5 |
| Gender |  |  |
| Man | 38.1 | 37.2 |
| Woman | 61.9 | 62.8 |
| Currently marital status |  |  |
| Never married | 7.2 | 7.4 |
| Currently married | 59.2 | 58.9 |
| Previously married | 33.6 | 33.7 |
| Completed education |  |  |
| No formal education | 8.5 | 7.9 |
| Primary | 73.1 | 72.8 |
| Secondary | 15.9 | 16.7 |
| Technical/University | 2.5 | 2.6 |
| Primary occupation |  |  |
| Agriculture or housework | 35.8 | 34.9 |
| Trading or shopkeeping | 20.7 | 21.0 |
| Bar work, waitressing, or sex work | 8.5 | 9.6 |
| Fishing-related occupation | 16.8 | 15.6 |
| Other | 18.2 | 18.9 |
| Religion |  |  |
| Catholic/Christian | 87.8 | 87.6 |
| Muslim | 11.6 | 11.8 |
| Other or none | 0.6 | 0.6 |
| Household wealth (quartile) |  |  |
| Lowest | 43.2 | 41.5 |
| Low-middle | 21.2 | 21.5 |
| High-middle | 21.8 | 22.2 |
| Highest | 13.6 | 14.8 |
| *Missing* | *0.2* | *n/a* |
| Migration |  |  |
| Long-term resident | 79.7 | 71.3 |
| In-migrant | 20.3 | 28.7 |
| Community type |  |  |
| Agrarian | 30.3 | 29.6 |
| Trading | 21.1 | 23.3 |
| Fishing | 48.6 | 47.0 |
| ***Behavioral*** |  |  |
| Number of sexual partners |  |  |
| 0–1 | 70.2 | 68.9 |
| ≥2 | 29.8 | 31.1 |
| Condom use |  |  |
| No partners or permanent partners only | 68.8 | 67.8 |
| Consistent use with casual partners | 13.7 | 13.8 |
| Inconsistent use with casual partners | 17.5 | 18.4 |
| Transactional sex |  |  |
| No | 48.6 | 47.3 |
| Yes | 51.4 | 52.7 |
| Any alcohol use consequences |  |  |
| No | 75.6 | 74.6 |
| Yes | 24.4 | 25.4 |
| Illicit drug use |  |  |
| No | 95.2 | 95.0 |
| Yes | 4.8 | 5.0 |
| ***HIV-related*** |  |  |
| ART use history (self-reported) |  |  |
| Never | 30.2 | 32.1 |
| Currently or previously | 69.8 | 67.9 |
| **HIV RNA viral load** |  |  |
| Viral load, in log_10_ copies/mL (*mean*) | 3.88 | 3.85 |
| <1,000 copies/mL | 74.4 | 73.1 |
| <400 copies/mL | 72.0 | 70.5 |
| <200 copies/mL | 71.1 | 69.6 |
| <50 copies/mL | 68.5 | 66.6 |

*^§^* Behavioral factors measured in the past year.

**Table S4.** Longitudinal virologic outcomes at visit-pair level, by viral load suppression (VLS) cutpoints (*N* = 4,604).

| **VLS Cutpoint** | **Durable VLS** | | |  | **New/Renewed VLS** | | |  | **Viral Rebound** | | |  | **Persistent Viremia** | | |
| --- | --- | --- | --- | --- | --- | --- | --- | --- | --- | --- | --- | --- | --- | --- | --- |
|  | n/N | Crude  % | Weighted  % |  | n/N | Crude  % | Weighted  % |  | n/N | Crude  % | Weighted  % |  | n/N | Crude  % | Weighted  % |
| <1,000 copies/mL | 3,536/4,604 | 76.8 | 75.3 |  | 531/4,604 | 11.5 | 12.3 |  | 108/4,604 | 2.4 | 2.5 |  | 429/4,604 | 9.3 | 9.9 |
| <400 copies/mL | 3,449/4,604 | 74.9 | 73.4 |  | 566/4,604 | 12.3 | 13.1 |  | 112/4,604 | 2.4 | 2.5 |  | 477/4,604 | 10.4 | 11.1 |
| <200 copies/mL | 3,408/4,604 | 74.0 | 72.4 |  | 575/4,604 | 12.5 | 13.3 |  | 113/4,604 | 2.5 | 2.5 |  | 508/4,604 | 11.0 | 11.8 |
| <50 copies/mL | 2,978/4,604 | 64.7 | 63.1 |  | 647/4,604 | 14.1 | 14.9 |  | 333/4,604 | 7.2 | 7.1 |  | 646/4,604 | 14.0 | 14.9 |

*Notes*: “Crude. %” represents the unweighted prevalence of each viral load outcome at visit-pair level, by survey attribute. “Weighted %” represents the prevalence of each viral load outcome at visit-pair level, corrected using stabilized inverse probability of selection weights.

**Figure S2**. Conditional probabilities of HIV viremia at follow-up in the visit-pair, by calendar period.

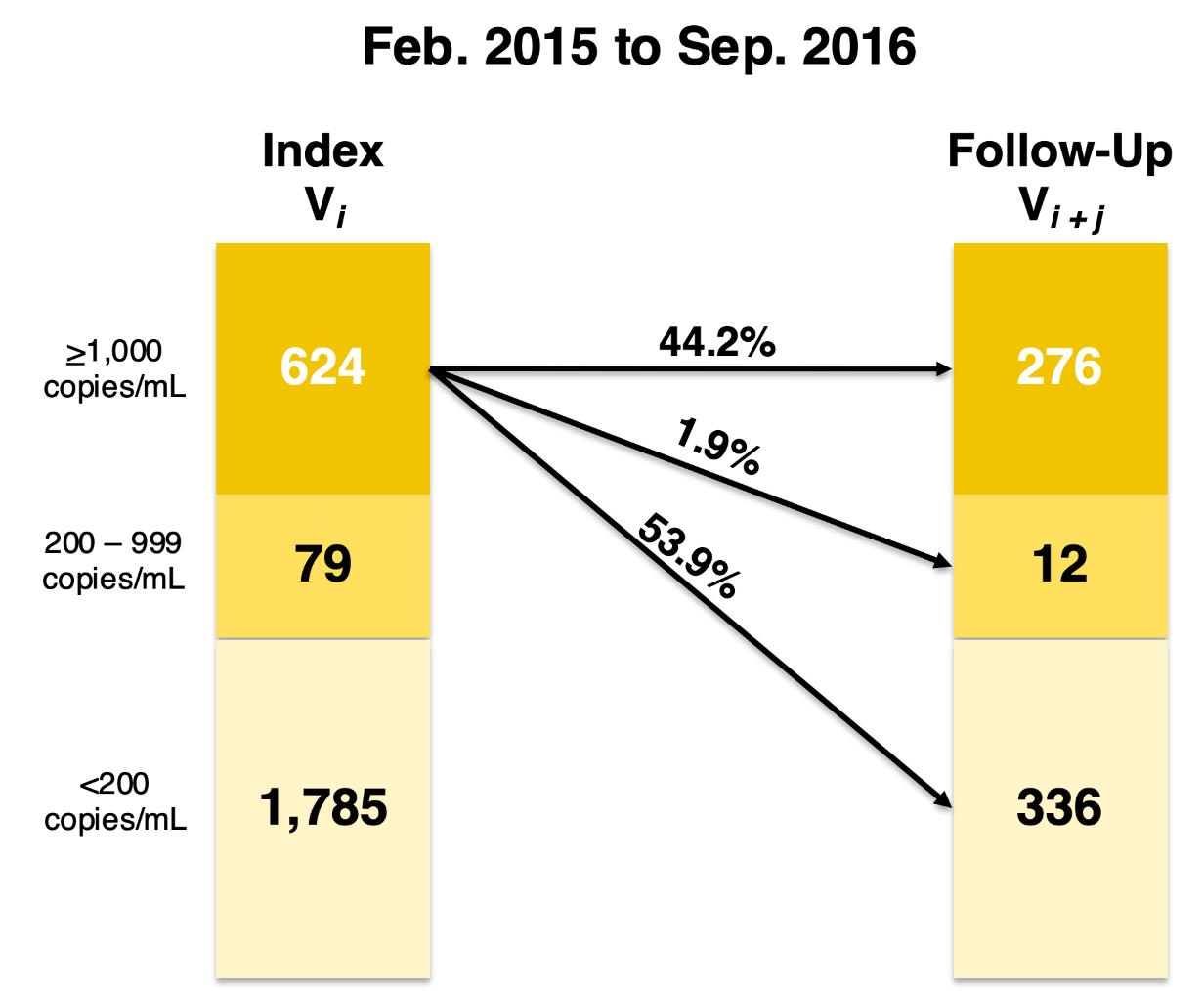

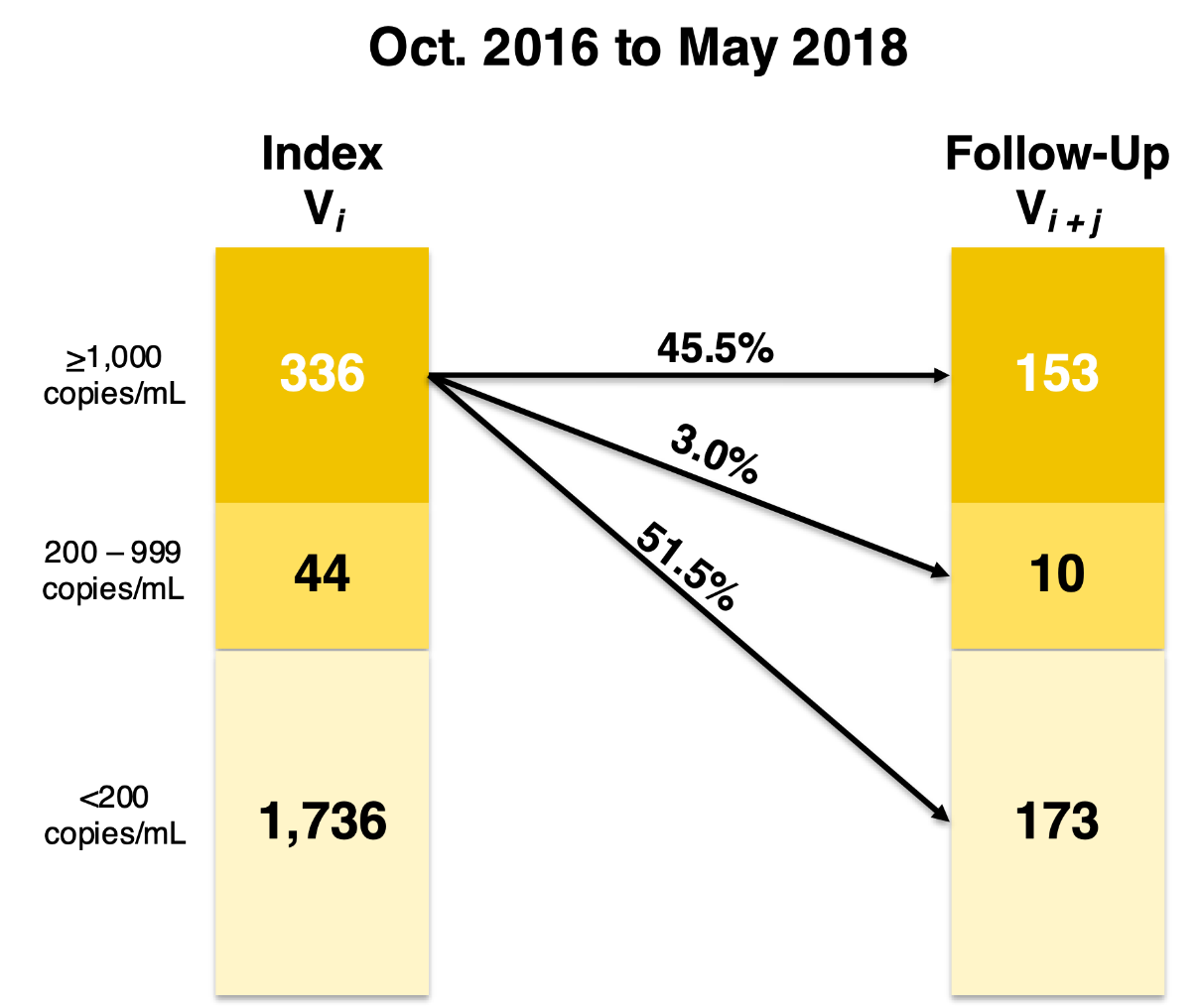

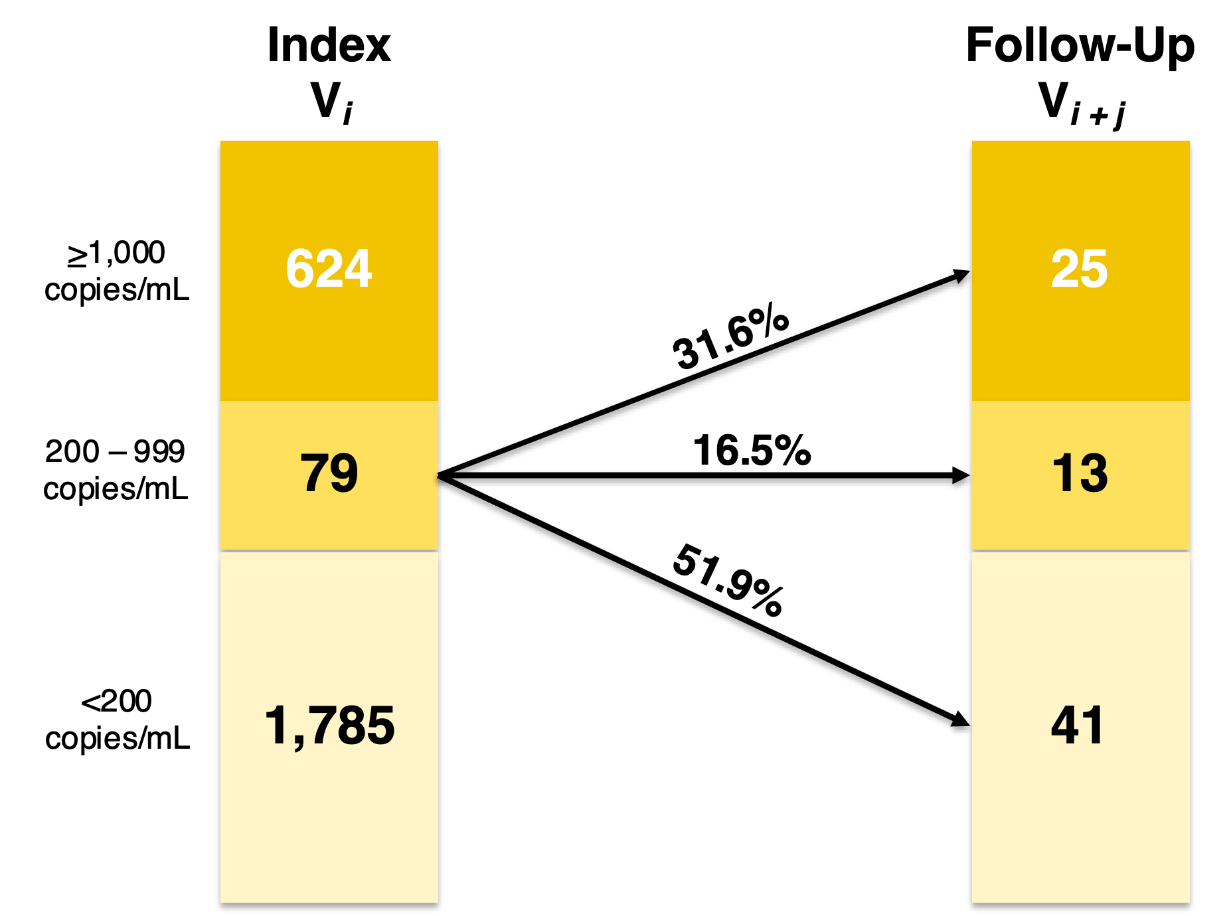

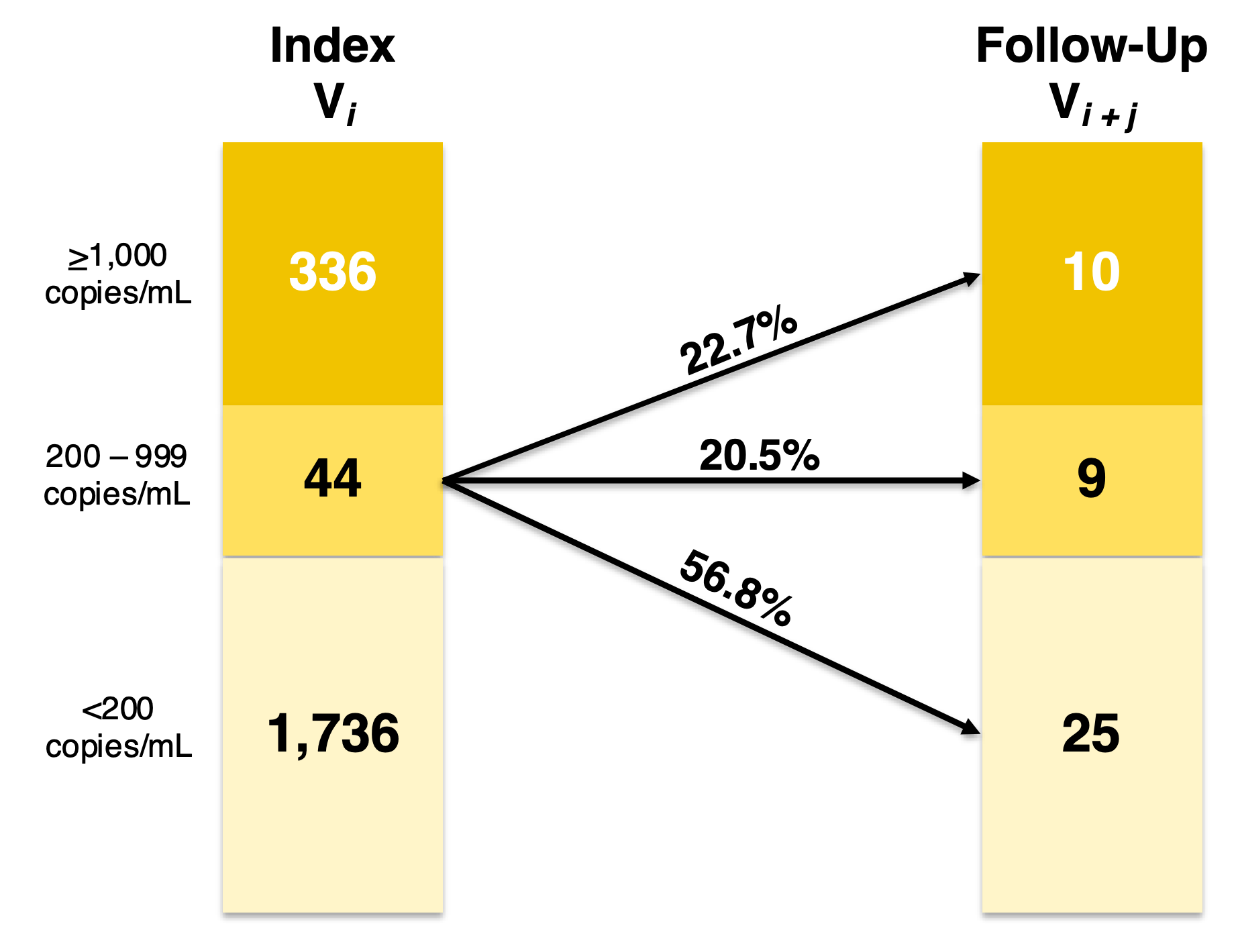

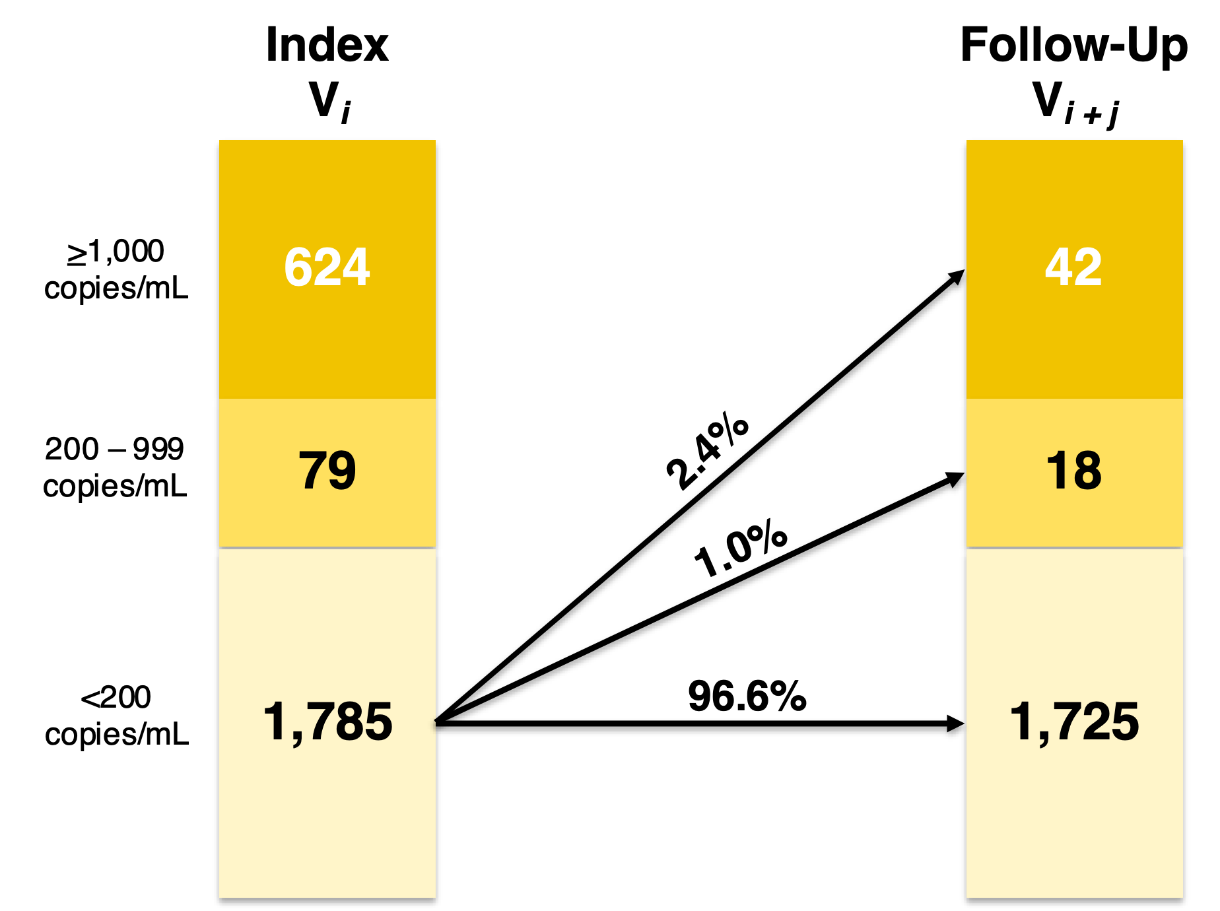

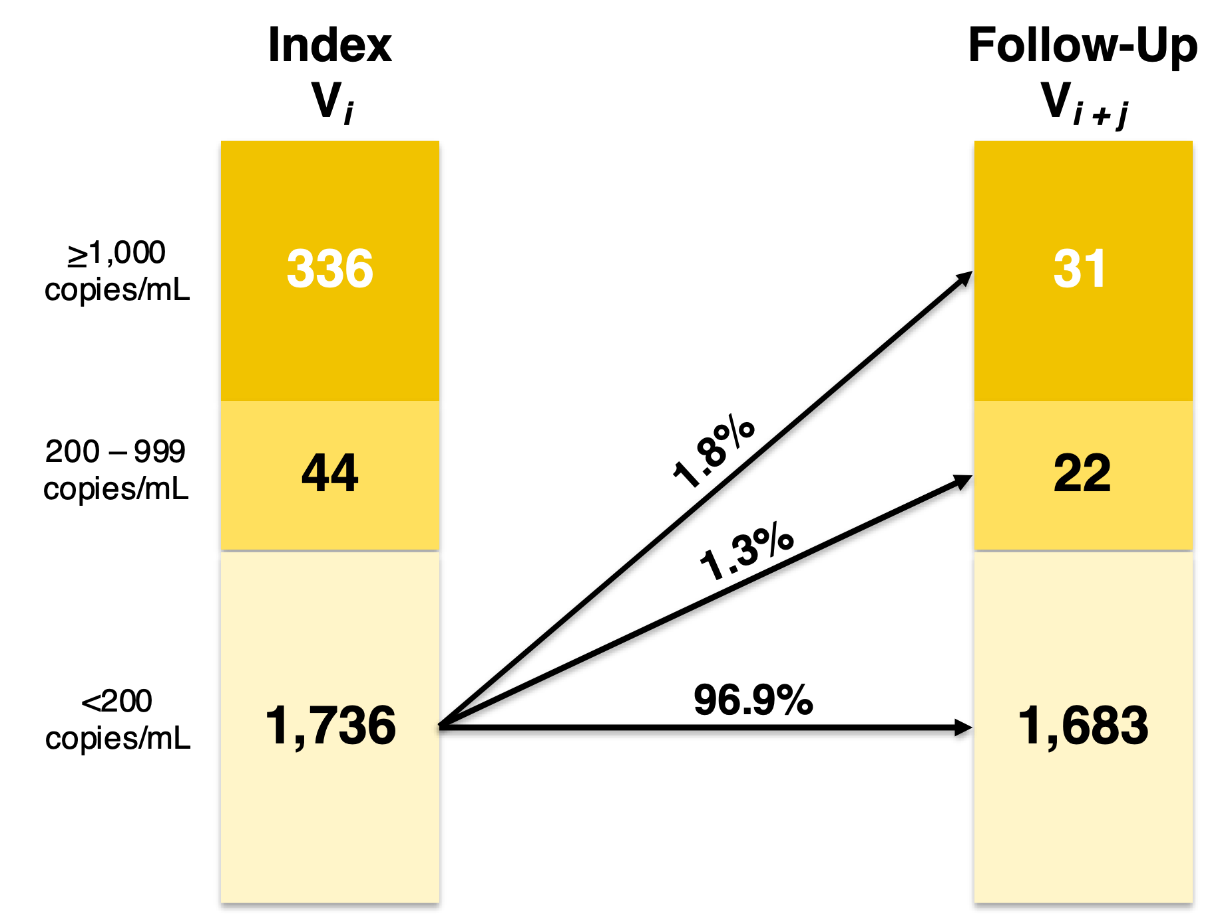

**Figure S3**. Conditional probabilities of HIV viremia at follow-up in the visit-pair, by community type.

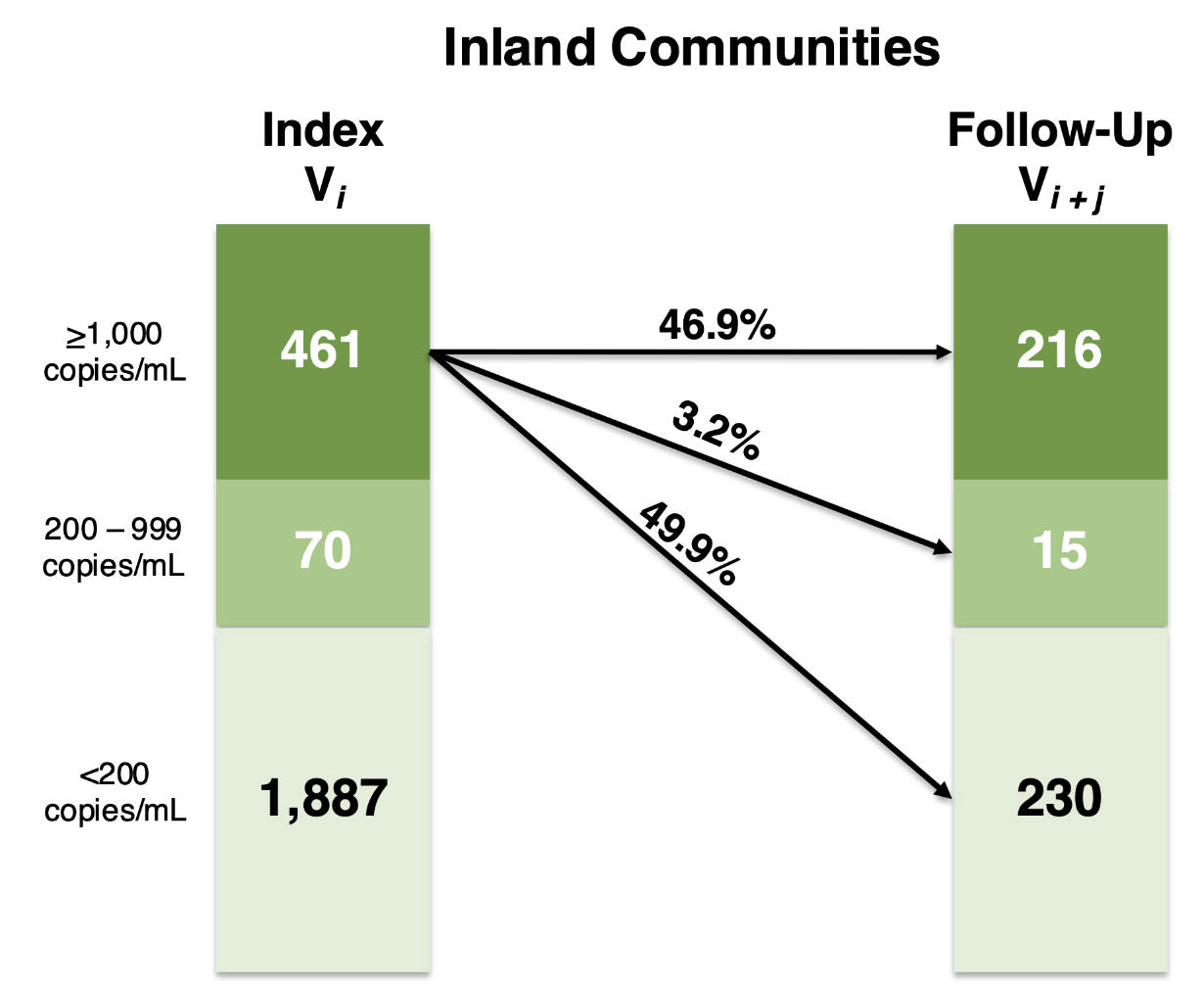

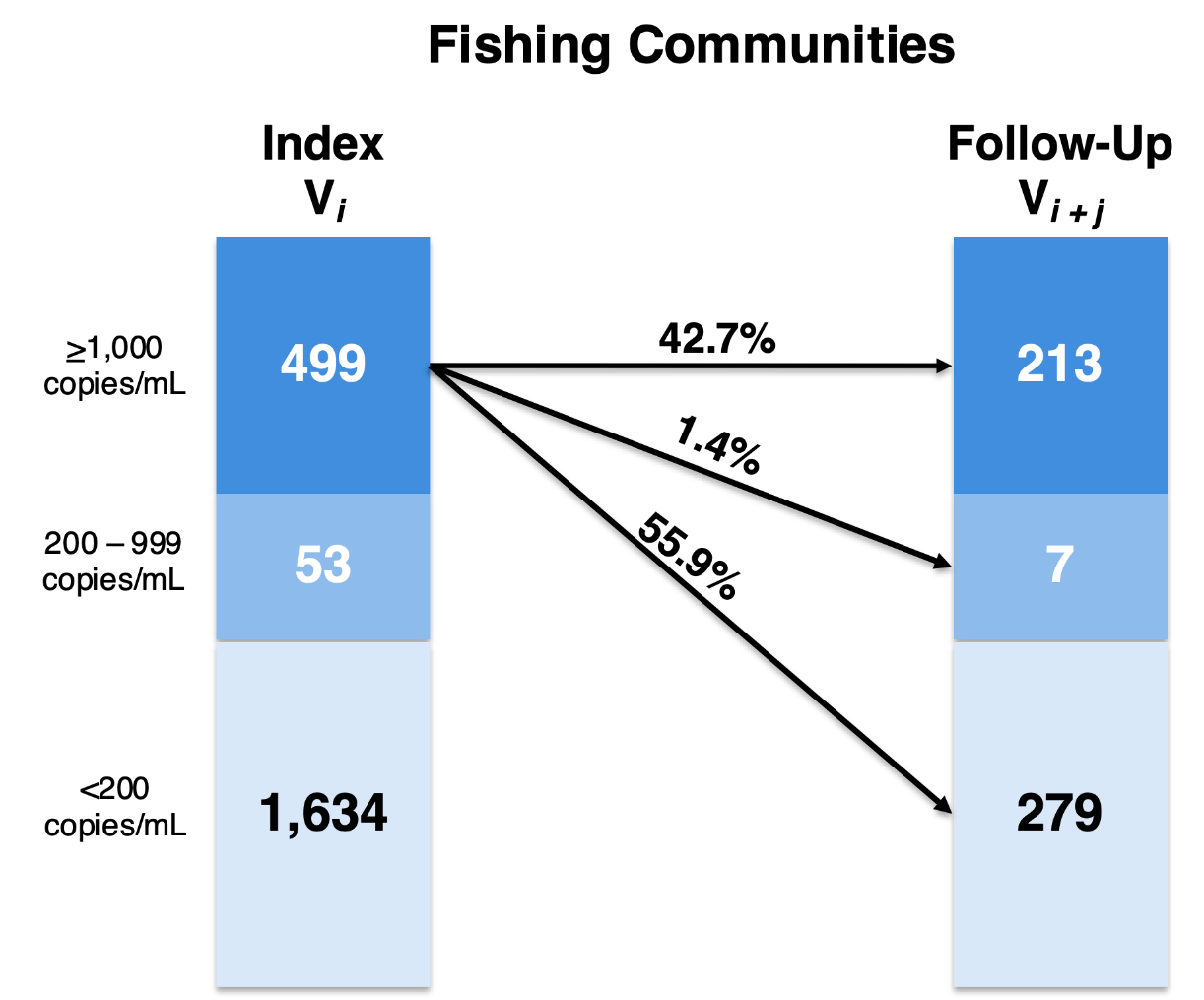

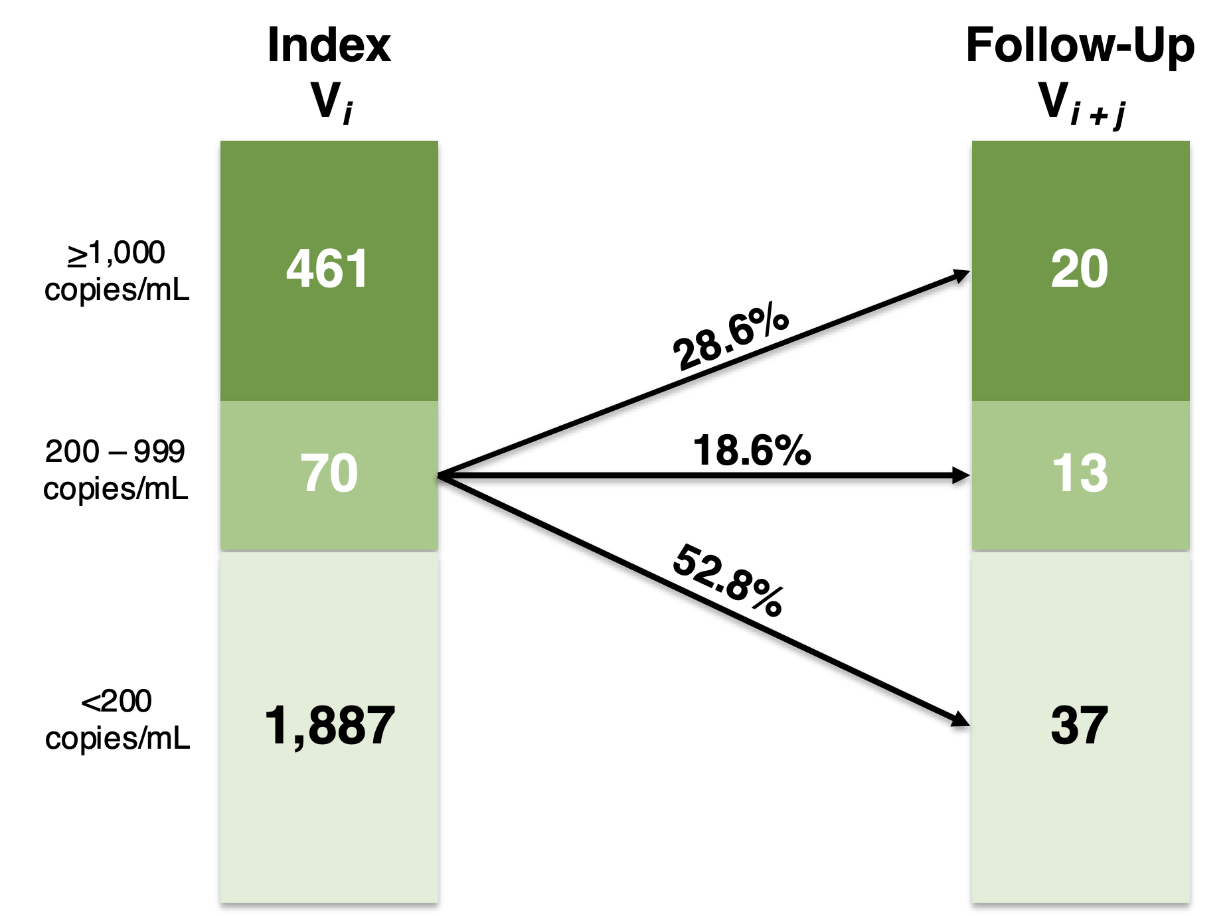

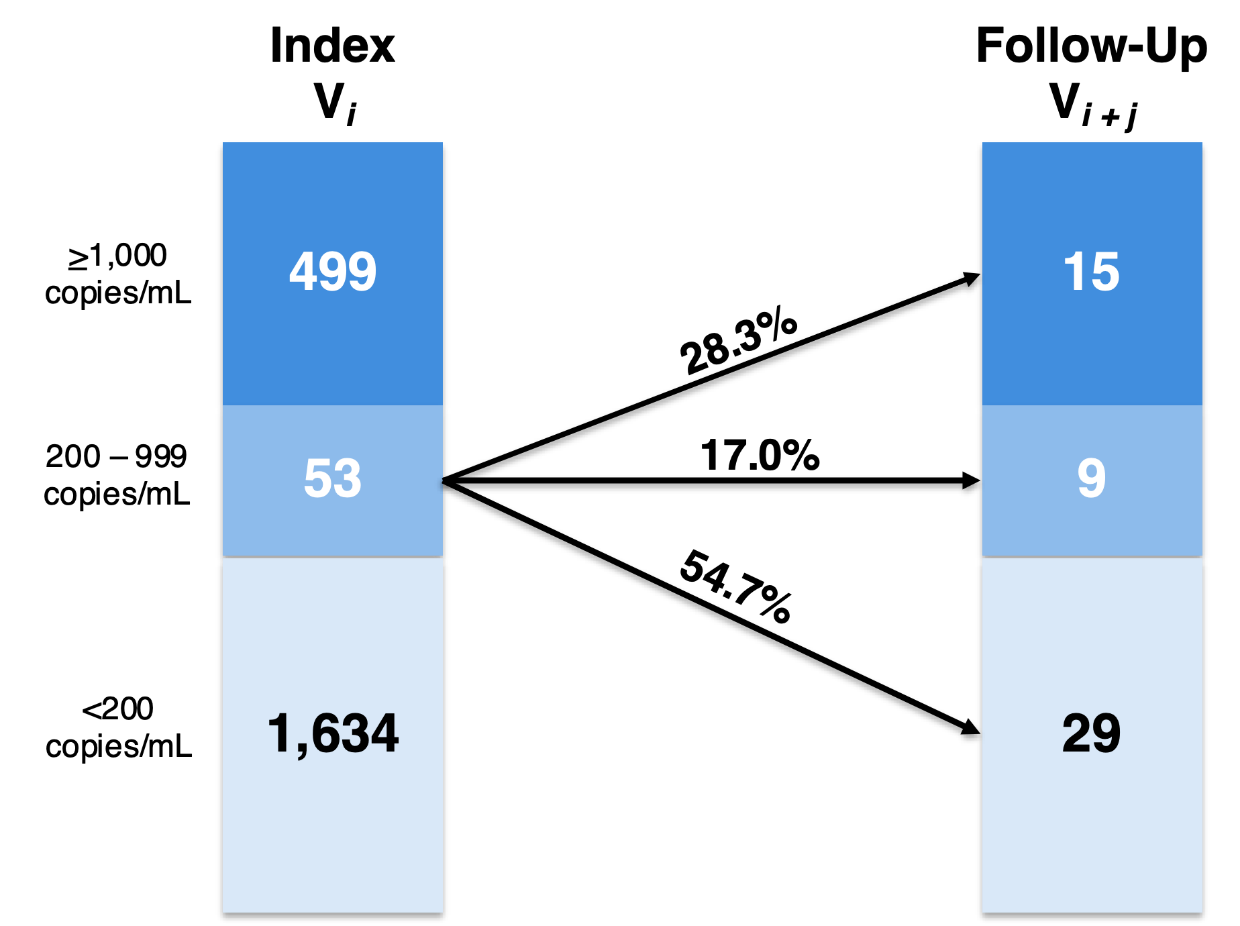

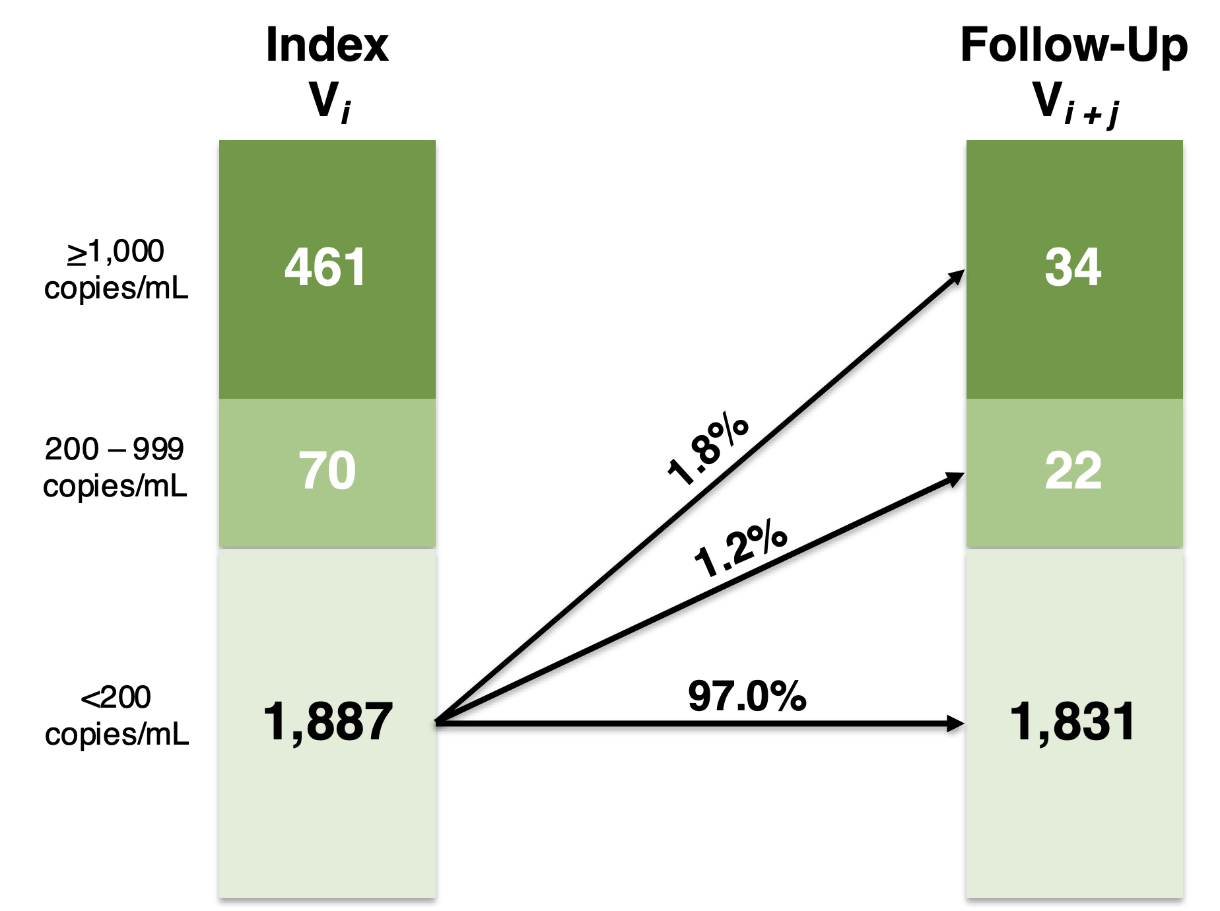

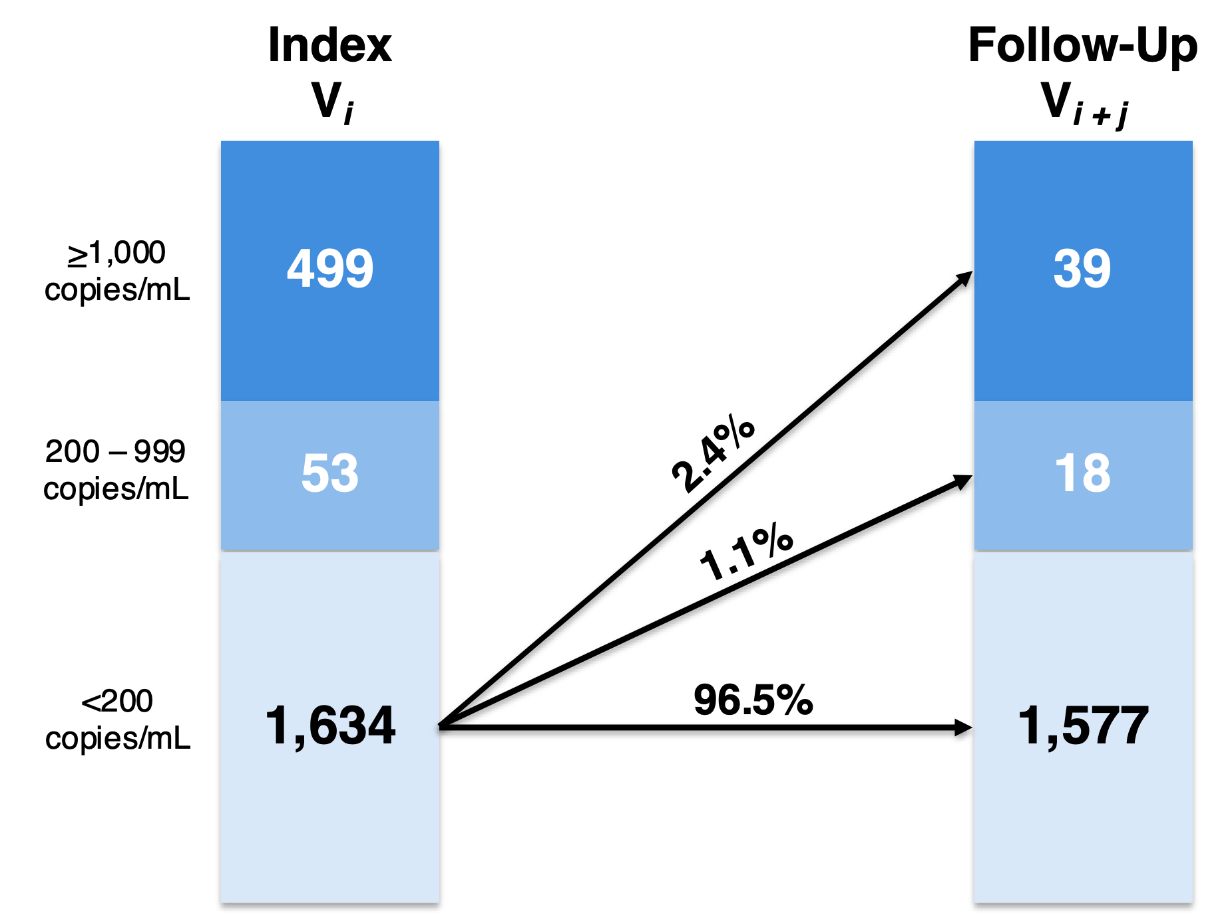

**Table S5**. Visit-pair-level characteristics of participants exhibiting persistent high-level viremia, by self-reported ART status.

| **Characteristics (*n*, %)** | **Total**  *N* = 429 | **ART <12 Months**  *n* = 340 (79.2%) | **ART ≥12 Months**  *n* = 89 (20.8%) | **χ²**  ***p*-value** |
| --- | --- | --- | --- | --- |
| ***Demographics*** |  |  |  |  |
| Index survey visit (calendar period) |  |  |  | **0.005** |
| Feb. 2015 – Sep. 2016 | 276 (64.3) | 230 (67.7) | 46 (51.7) |  |
| Oct. 2016 – May 2018 | 153 (35.7) | 110 (32.3) | 43 (48.3) |  |
| Age, in years (*median*, *IQR*)† | 31 (27–35) | 30 (26–35) | 33 (28–36) | **0.033** |
| Age group |  |  |  | 0.080 |
| 15-29 years | 190 (44.3) | 159 (46.8) | 31 (34.8) |  |
| 30-39 years | 174 (40.6) | 129 (37.9) | 45 (50.6) |  |
| 40-49 years | 65 (15.1) | 52 (15.3) | 13 (14.6) |  |
| Gender |  |  |  | **0.001** |
| Man | 245 (57.1) | 208 (61.2) | 37 (41.6) |  |
| Woman | 184 (42.9) | 132 (38.8) | 52 (58.4) |  |
| Currently marital status |  |  |  | 0.505 |
| Never married | 53 (12.4) | 45 (13.2) | 8 (9.0) |  |
| Currently married | 243 (56.6) | 189 (55.6) | 54 (60.7) |  |
| Previously married | 133 (31.0) | 106 (31.2) | 27 (30.3) |  |
| Completed education |  |  |  | 0.310 |
| No formal education | 27 (6.3) | 20 (5.9) | 17 (7.9) |  |
| Primary | 304 (70.9) | 237 (69.7) | 67 (75.3) |  |
| Secondary | 91 (21.2) | 76 (22.3) | 15 (16.8) |  |
| Technical/University | 7 (1.6) | 7 (2.1) | *n/a* |  |
| Primary occupation |  |  |  | 0.106 |
| Agriculture or housework | 135 (31.5) | 108 (31.8) | 27 (30.3) |  |
| Trading or shopkeeping | 83 (19.3) | 66 (19.4) | 17 (19.1) |  |
| Bar work, waitressing, or sex work | 20 (4.7) | 11 (3.2) | 9 (10.1) |  |
| Fishing-related occupation | 107 (24.9) | 87 (25.6) | 20 (22.5) |  |
| Other | 84 (19.6) | 68 (20.0) | 16 (18.0) |  |
| Religion |  |  |  | 0.404 |
| Catholic/Christian | 379 (87.3) | 297 (87.3) | 82 (92.1) |  |
| Muslim | 48 (11.2) | 41 (12.1) | 7 (7.9) |  |
| Other/none | 2 (0.5) | 2 (0.6) | *n/a* |  |
| Household wealth (quartile) |  |  |  | 0.360 |
| Lowest | 202 (47.1) | 156 (45.9) | 146 (51.7) |  |
| Low-middle | 85 (19.8) | 74 (21.8) | 11 (12.4) |  |
| High-middle | 98 (22.8) | 75 (22.0) | 23 (25.8) |  |
| Highest | 43 (10.0) | 34 (10.0) | 9 (10.1) |  |
| *Missing* | *1 (0.2)* | *1 (0.3)* | *n/a* |  |
| Migration |  |  |  | 0.506 |
| Long-term resident | 341 (79.5) | 268 (78.8) | 73 (82.0) |  |
| In-migrant | 88 (20.5) | 72 (21.2) | 16 (18.0) |  |
| Community type |  |  |  | 0.142 |
| Agrarian | 129 (30.1) | 103 (30.3) | 26 (29.2) |  |
| Trading | 87 (20.3) | 75 (22.1) | 12 (13.5) |  |
| Fishing | 213 (49.6) | 162 (47.6) | 51 (57.3) |  |
| ***Behavioral*** |  |  |  |  |
| Number of sexual partners |  |  |  | 0.496 |
| 0–1 | 276 (64.3) | 216 (63.5) | 60 (67.4) |  |
| ≥2 | 153 (35.7) | 124 (36.5) | 29 (32.6) |  |
| Condom use |  |  |  | 0.267 |
| No partners or permanent partners only | 273 (63.6) | 211 (62.1) | 62 (69.7) |  |
| Consistent use with casual partners | 73 (17.0) | 58 (17.0) | 15 (16.8) |  |
| Inconsistent use with casual partners | 83 (19.4) | 71 (20.9) | 12 (13.5) |  |
| Transactional sex |  |  |  | 0.193 |
| No | 243 (56.6) | 198 (58.2) | 45 (50.6) |  |
| Yes | 186 (43.4) | 142 (41.8) | 44 (49.4) |  |
| Any alcohol use consequences |  |  |  | **0.021** |
| No | 321 (74.8) | 246 (72.3) | 75 (84.3) |  |
| Yes | 108 (25.2) | 94 (27.6) | 15 (15.7) |  |
| Illicit drug use |  |  |  | 0.487 |
| No | 403 (93.9) | 318 (93.5) | 85 (95.5) |  |
| Yes | 26 (6.1) | 22 (6.5) | 4 (4.5) |  |
| Intimate partner violence |  |  |  | 0.885 |
| No | 301 (70.2) | 238 (70.0) | 63 (70.8) |  |
| Yes | 128 (29.8) | 102 (30.0) | 26 (29.2) |  |

*Notes*: Individuals on ART <1 year also included those reporting never or previous ART use throughout the observation period. † *p*-values calculated using Wilcoxon rank-sum tests comparing median values with interquartile ranges (IQR). *^§^* Behavioral factors measured in the past year.

**Table S6**. Visit-pair-level characteristics of participants self-reporting ART use for ≥12 months, by durable viral load suppression (VLS) and persistent high-level viremia.

| **Characteristics (*n*, %)** | **Total**  *N* = 3,234 | **Durable VLS**  *n* = 3,145 (97.2%) | **Persistent Viremia**  *n* = 89 (2.8%) | **χ²**  ***p*-value** |
| --- | --- | --- | --- | --- |
| ***Demographics*** |  |  |  |  |
| Index survey visit (calendar period) |  |  |  | 0.664 |
| Feb. 2015 – Sep. 2016 | 1,598 | 1,552 (97.1) | 46 (2.9) |  |
| Oct. 2016 – May 2018 | 1,636 | 1,593 (97.4) | 43 (2.6) |  |
| Age, in years (*median*, *IQR*)† | 36 (31–41) | 36 (31–41) | 33 (28–36) | **<0.001** |
| Age group |  |  |  | **<0.001** |
| 15-29 years | 641 | 610 (95.2) | 31 (4.8) |  |
| 30-39 years | 1,557 | 1,512 (97.1) | 45 (2.9) |  |
| 40-49 years | 1,036 | 1,023 (98.7) | 13 (1.3) |  |
| Gender |  |  |  | 0.071 |
| Man | 1,058 | 1,021 (96.5) | 37 (3.5) |  |
| Woman | 2,176 | 2,124 (97.6) | 52 (2.4) |  |
| Currently marital status |  |  |  | 0.320 |
| Never married | 178 | 170 (95.5) | 8 (4.5) |  |
| Currently married | 1,976 | 1,922 (97.3) | 54 (2.7) |  |
| Previously married | 1,080 | 1,053 (97.5) | 27 (2.5) |  |
| Completed education |  |  |  | 0.477 |
| No formal education | 293 | 286 (97.6) | 7 (2.4) |  |
| Primary | 2,349 | 2,282 (97.1) | 67 (2.9) |  |
| Secondary | 514 | 499 (97.1) | 15 (2.9) |  |
| Technical/University | 78 | 78 (100) | *n/a* |  |
| Primary occupation |  |  |  | 0.149 |
| Agriculture or housework | 1,261 | 1,234 (97.9) | 27 (2.1) |  |
| Trading or shopkeeping | 672 | 655 (97.5) | 17 (2.5) |  |
| Bar work, waitressing, or sex work | 274 | 265 (96.7) | 9 (3.3) |  |
| Fishing-related occupation | 455 | 435 (95.6) | 20 (4.4) |  |
| Other | 572 | 556 (97.2) | 16 (2.8) |  |
| Religion |  |  |  | 0.444 |
| Catholic/Christian | 2,857 | 2,775 (97.1) | 82 (2.9) |  |
| Muslim | 352 | 345 (98.0) | 7 (2.0) |  |
| Other/none | 25 | 25 (100) | *n/a* |  |
| Household wealth (quartile) |  |  |  | 0.175 |
| Lowest | 1,350 | 1,304 (96.6) | 46 (3.4) |  |
| Low-middle | 620 | 609 (98.2) | 11 (1.8) |  |
| High-middle | 762 | 739 (97.0) | 23 (3.0) |  |
| Highest | 499 | 490 (98.2) | 9 (1.8) |  |
| *Missing* | *3* | *3 (100)* | *n/a* |  |
| Migration |  |  |  | **0.019** |
| Long-term resident | 2,895 | 2,822 (97.5) | 73 (2.5) |  |
| In-migrant | 339 | 323 (95.3) | 16 (4.7) |  |
| Community type |  |  |  | 0.085 |
| Agrarian | 1,037 | 1,011 (97.5) | 26 (2.5) |  |
| Trading | 684 | 627 (98.3) | 12 (1.7) |  |
| Fishing | 1,513 | 1,462 (96.6) | 51 (3.4) |  |
| ***Behavioral*** |  |  |  |  |
| Number of sexual partners |  |  |  | **0.021** |
| 0–1 | 2,505 | 2,445 (97.6) | 60 (2.4) |  |
| ≥2 | 729 | 700 (96.0) | 29 (4.0) |  |
| Condom use |  |  |  | 0.266 |
| No partners or permanent partners only | 2,452 | 2,390 (97.5) | 62 (2.5) |  |
| Consistent use with casual partners | 377 | 362 (96.0) | 15 (4.0) |  |
| Inconsistent use with casual partners | 405 | 393 (97.0) | 12 (3.0) |  |
| Transactional sex |  |  |  | 0.804 |
| No | 1,677 | 1,632 (97.3) | 45 (2.7) |  |
| Yes | 1,557 | 1,513 (97.2) | 44 (2.8) |  |
| Any alcohol use consequences |  |  |  | 0.523 |
| No | 2,799 | 2,724 (97.3) | 75 (2.7) |  |
| Yes | 435 | 421 (96.8) | 14 (3.2) |  |
| Illicit drug use |  |  |  | 0.501 |
| No | 3,129 | 3,044 (97.3) | 85 (2.7) |  |
| Yes | 105 | 101 (96.2) | 4 (3.8) |  |
| Intimate partner violence |  |  |  | 0.304 |
| No | 2,439 | 2,376 (97.4) | 63 (2.6) |  |
| Yes | 795 | 769 (96.7) | 26 (3.3) |  |

*Notes*: Durable VLS was defined as consecutive visits with <200 HIV RNA copies/mL. Persistent high-level viremia was defined as consecutive visits with >1,000 HIV RNA copies/mL. † *p*-values calculated using Wilcoxon rank-sum tests comparing median values with interquartile ranges (IQR). *^§^* Behavioral factors measured in the past year.

**Table S7**. Weighted region-level prevalence of persistent high-level viremia, by gender and calendar period.

| **Region** | **Survey Interval 1:**  Jun. 2015 to Sep. 2016 | | |  | **Survey Interval 2:**  Oct. 2016 to May 2018 | | |  | **Mann-Whitney U Test**  % Diff. (*p*-value)* | | |
| --- | --- | --- | --- | --- | --- | --- | --- | --- | --- | --- | --- |
|  | Men  *n* = 930 | Women  *n* = 1,558 | Total  *N* = 2,488 |  | Men  *n* = 779 | Women  *n* = 1,337 | Total  *N* = 2,116 |  | Men | Women | Total |
| Ddimo | 21.5  (16.4–26.7) | 7.7  (4.9–10.5) | 13.3  (10.6–16.0) |  | 12.2  (7.7–16.6) | 7.5  (4.6–10.4) | 9.3  (6.8–11.8) |  | **–9.3**  **(0.010)** | –0.2  (0.567) | **–4.0**  **(0.015)** |
| Kabira | 38.0  (22.9–53.0) | 12.7  (6.0–19.4) | 20.3  (13.5–27.0) |  | 13.6  (2.0–25.1) | 9.7  (3.2–16.3) | 10.9  (5.1–16.6) |  | **–24.4**  **(0.048)** | –3.0  (0.604) | –9.4  (0.094) |
| Kakuuto | 5.8  (<0.1–14.0) | 11.0  (4.8–17.3) | 9.7  (4.6–14.8) |  | 15.0  (1.3–28.8) | 3.2  (<0.1–7.3) | 6.2  (1.4–11.0) |  | 9.2  (0.485) | –7.8  (0.094) | –3.5  (0.324) |
| Kalisizo | 10.3  (3.1–17.5) | 4.7  (0.8–8.5) | 6.8  (3.2–10.4) |  | 11.8  (3.4–20.3) | 5.6  (1.2–10.0) | 7.7  (3.7–11.9) |  | 1.5  (0.949) | 0.9  (0.839) | 0.9  (0.891) |
| Kasasa/Sanje | 17.2  (8.5–26.0) | 5.3  (2.1–8.5) | 8.5  (5.2–11.9) |  | 16.6  (8.1–25.1) | 4.4  (14.8–7.2) | 7.8  (4.6–11.0) |  | –0.6  (0.767) | –0.9  (0.444) | –0.7  (0.480) |
| Kasensero | 16.6  (12.5–20.6) | 8.2  (5.5–11.0) | 12.0  (9.6–14.4) |  | 8.6  (5.4–11.8) | 5.9  (3.4–8.5) | 7.2  (5.1–9.2) |  | **–8.0**  **(0.004)** | –2.3  (0.176) | **–4.8**  **(0.002)** |
| Kibaale/Rakai | 11.3  (<0.1–22.8) | 1.9  (<0.1–5.6) | 5.2  (0.5–10.0) |  | 10.0  (<0.1–29.6) | 2.2  (<0.1–6.5) | 3.5  (<0.1–8.4) |  | –1.3  (1.000) | –0.3  (0.899) | –1.7  (0.745) |
| Kyotera | 15.0  (4.1–26.0) | 9.9  (4.8–15.2) | 11.2  (6.5–16.0) |  | 29.0  (9.6–48.4) | 6.5  (1.1–12.0) | 11.6  (5.3–17.8) |  | 14.0  (0.399) | –3.4  (0.251) | 0.4  (0.638) |
| Lwanda | 18.7  (8.7–28.6) | 10.3  (4.8–15.9) | 13.2  (8.2–18.2) |  | 11.9  (2.8–20.9) | 5.2  (0.8–9.6) | 7.5  (3.3–11.7) |  | –6.8  (0.219) | 1.6  (0.119) | **–5.7**  **(0.048)** |
| Lyantonde | 26.4  (<0.1–76.4) | *n/a* | 4.2  (<0.1–12.3) |  | *n/a* | *n/a* | *n/a* |  | *n/a* | *n/a* | *n/a* |

***Bolded** values represent statistically significant region-level median differences in persistent high-level viremia over time at the *p* < 0.05 level or below.

**Table S8**. Weighted community-level prevalence of persistent high-level viremia, by calendar period.

| **Geo-masked**  **Community** |  | **Survey Interval 1:**  Jun. 2015 to Sep. 2016 | | | | |  | **Survey Interval 2:**  Oct. 2016 to May 2018 | | | | |  | **Mann-Whitney**  **U Test** | |
| --- | --- | --- | --- | --- | --- | --- | --- | --- | --- | --- | --- | --- | --- | --- | --- |
|  |  | *N* | Weighted % | 95% CI | | |  | *N* | Weighted % | 95% CI | | |  | % Diff. | *p*-value* |
| 1 |  | 13 | 8.1 | <0.1 | to | 23.6 |  | *n/a* | *n/a* | *n/a* | | |  | *n/a* | *n/a* |
| 2 |  | 52 | 10.1 | 1.8 | to | 18.4 |  | 40 | 6.9 | <0.1 | to | 14.9 |  | –3.2 | 0.606 |
| 4 |  | 45 | 12.8 | 2.9 | to | 22.7 |  | 28 | 8.2 | <0.1 | to | 18.6 |  | –4.6 | 0.578 |
| 5 |  | 43 | 8.1 | <0.1 | to | 16.4 |  | 35 | 4.2 | <0.1 | to | 11.0 |  | –3.9 | 0.415 |
| 6 |  | 45 | 6.9 | <0.1 | to | 14.4 |  | 37 | 15.8 | 3.9 | to | 27.8 |  | 8.9 | 0.507 |
| 7 |  | 47 | 14.2 | 4.1 | to | 24.3 |  | 37 | 9.9 | 0.2 | to | 19.7 |  | –4.3 | 0.785 |
| 8 |  | 30 | 21.9 | 6.8 | to | 37.0 |  | 28 | 9.2 | <0.1 | to | 20.2 |  | –12.7 | 0.208 |
| 16 |  | 19 | 14.5 | <0.1 | to | 30.8 |  | 16 | 10.6 | <0.1 | to | 26.2 |  | –3.9 | 0.785 |
| 19 |  | 54 | 14.8 | 5.2 | to | 24.4 |  | 42 | 9.7 | 0.7 | to | 18.8 |  | –5.1 | 0.244 |
| 23 |  | 56 | 10.5 | 2.4 | to | 18.6 |  | 56 | 4.8 | <0.1 | to | 10.5 |  | –5.7 | 0.299 |
| 24 |  | 73 | 9.6 | 2.8 | to | 16.4 |  | 47 | 6.8 | <0.1 | to | 14.2 |  | –2.8 | 0.537 |
| 29 |  | 31 | 3.3 | <0.1 | to | 9.6 |  | 28 | 7.3 | <0.1 | to | 17.1 |  | 4.0 | 0.498 |
| 33 |  | 29 | 8.4 | <0.1 | to | 18.7 |  | 21 | 0 | *n/a* | | |  | –8.4 | 0.224 |
| 34 |  | 73 | 6.6 | 0.9 | to | 12.3 |  | 65 | 10.8 | 3.2 | to | 18.4 |  | 4.2 | 0.608 |
| 36 |  | 31 | 3.3 | <0.1 | to | 9.7 |  | 22 | 0 | *n/a* | | |  | –3.3 | 0.400 |
| 38 |  | 609 | 11.9 | 9.4 | to | 14.5 |  | 549 | 7.3 | 5.2 | to | 9.5 |  | **–4.6** | **0.007** |
| 40 |  | 50 | 7.7 | 0.2 | to | 15.2 |  | 51 | 10.2 | 1.8 | to | 18.6 |  | 2.5 | 0.752 |
| 55 |  | 38 | 8.9 | <0.1 | to | 18.1 |  | 20 | 10.0 | <0.1 | to | 23.5 |  | 1.1 | 0.788 |
| 57 |  | 24 | 9.4 | <0.1 | to | 21.4 |  | 21 | 19.0 | 1.8 | to | 36.2 |  | 9.6 | 0.531 |
| 58 |  | 13 | 17.9 | <0.1 | to | 39.7 |  | 15 | 14.1 | <0.1 | to | 32.3 |  | –3.8 | 0.879 |
| 62 |  | 36 | 3.0 | <0.1 | to | 8.6 |  | 31 | 0 | *n/a* | | |  | –3.0 | 0.353 |
| 74 |  | 10 | 20.2 | <0.1 | to | 46.5 |  | 11 | 24.5 | <0.1 | to | 51.1 |  | 4.3 | 0.918 |
| 77 |  | 23 | 17.8 | 1.8 | to | 33.8 |  | 26 | 7.3 | <0.1 | to | 17.4 |  | –10.5 | 0.306 |
| 89 |  | 28 | 6.6 | <0.1 | to | 16.0 |  | 26 | 9.2 | <0.1 | to | 20.5 |  | 2.6 | 0.939 |
| 106 |  | 60 | 24.4 | 13.4 | to | 35.4 |  | 50 | 13.0 | 3.6 | to | 22.4 |  | –11.4 | 0.184 |
| 107 |  | 31 | 7.4 | <0.1 | to | 16.8 |  | 22 | 7.3 | <0.1 | to | 18.4 |  | –0.1 | 0.770 |
| 108 |  | 46 | 6.6 | <0.1 | to | 13.9 |  | 57 | 7.4 | 0.5 | to | 14.3 |  | 0.8 | 0.787 |
| 120 |  | 9 | 0 | *n/a* | | |  | 8 | 18.7 | <0.1 | to | 47.6 |  | 18.7 | 0.289 |
| 370 |  | 43 | 15.4 | 4.5 | to | 26.4 |  | 27 | 8.1 | <0.1 | to | 18.5 |  | –7.3 | 0.406 |
| 391 |  | 27 | 22.8 | 6.7 | to | 39.0 |  | 25 | 0 | *n/a* | | |  | **–22.8** | **0.013** |
| 602 |  | 99 | 13.2 | 6.5 | to | 19.9 |  | 88 | 3.4 | <0.1 | to | 7.2 |  | **–9.8** | **0.031** |
| 754 |  | 42 | 10.4 | 1.1 | to | 19.8 |  | 16 | 21.3 | 0.5 | to | 42.0 |  | 10.9 | 0.742 |
| 770 |  | 127 | 11.5 | 5.9 | to | 17.1 |  | 132 | 9.8 | 4.7 | to | 14.9 |  | –1.7 | 0.606 |
| 771 |  | 302 | 11.8 | 8.1 | to | 15.4 |  | 246 | 6.1 | 3.1 | to | 9.1 |  | **–5.7** | **0.027** |
| 772 |  | 26 | 15.5 | 1.3 | to | 29.6 |  | 20 | 4.1 | <0.1 | to | 13.1 |  | –11.4 | 0.267 |
| 773 |  | 31 | 23.0 | 7.9 | to | 38.1 |  | 27 | 28.9 | 11.5 | to | 46.4 |  | 5.9 | 0.790 |
| 774 |  | 117 | 15.9 | 9.2 | to | 22.6 |  | 102 | 11.0 | 4.9 | to | 17.1 |  | –4.9 | 0.194 |

***Bolded** values represent statistically significant community-level median differences in persistent high-level viremia over time at the *p* < 0.05 level or below.

**Table S9**. Descriptive sample statistics at the index visit for participants contributing one visit-pair versus two visit-pairs to the analysis—2015 to 2020.

| **Characteristics (*n*, %)** | **1 Visit-Pair**  *n* = 1,556 (50.5%) | **2 Visit-Pairs**  *n* = 1,524 (49.5%) | **Total**  *N* = 3,080 | **χ²**  ***p*-value***** |
| --- | --- | --- | --- | --- |
| ***Demographics*** |  |  |  |  |
| Age, in years (*median*, *IQR*)† | 33 (28–39) | 35 (30–40) | 34 (28–39) | **<0.001** |
| Age group |  |  |  | **<0.001** |
| 15-29 years | 528 (33.9) | 380 (24.9) | 908 (29.5) |  |
| 30-39 years | 661 (42.5) | 760 (49.9) | 1,421 (46.1) |  |
| 40-49 years | 367 (23.6) | 384 (25.2) | 751 (24.4) |  |
| Gender |  |  |  | **0.001** |
| Man | 637 (40.9) | 536 (35.2) | 1,173 (38.1) |  |
| Woman | 919 (59.1) | 988 (64.8) | 1,907 (61.9) |  |
| Currently marital status |  |  |  | **0.007** |
| Never married | 133 (8.5) | 88 (5.8) | 221 (7.2) |  |
| Currently married | 894 (57.5) | 929 (61.0) | 1,823 (60.0) |  |
| Previously married | 529 (34.0) | 507 (33.3) | 1,036 (33.6) |  |
| Educational attainment |  |  |  | 0.730 |
| No formal education | 132 (8.5) | 128 (8.4) | 260 (8.5) |  |
| Primary | 1,140 (73.2) | 1,112 (73.0) | 2,252 (73.1) |  |
| Secondary | 241 (15.5) | 250 (16.4) | 491 (15.9) |  |
| Technical/University | 43 (2.8) | 23 (2.2) | 77 (2.5) |  |
| Primary occupation |  |  |  | **0.004** |
| Agriculture or housework | 511 (32.8) | 590 (38.7) | 1,101 (35.8) |  |
| Trading or shopkeeping | 323 (20.8) | 314 (20.6) | 637 (20.7) |  |
| Bar work, waitressing, or sex work | 136 (8.7) | 125 (8.2) | 261 (8.5) |  |
| Fishing-related occupation | 292 (18.8) | 227 (14.9) | 519 (16.8) |  |
| Other | 294 (18.9) | 268 (17.6) | 562 (18.2) |  |
| Religion |  |  |  | 0.260 |
| Catholic/Christian | 1,356 (87.2) | 1,348 (88.4) | 2,704 (87.8) |  |
| Muslim | 192 (12.3) | 164 (10.8) | 356 (11.6) |  |
| Other/none | 8 (0.5) | 12 (0.8) | 20 (0.6) |  |
| Household wealth (quartile) |  |  |  | **0.048** |
| Lowest | 685 (44.0) | 646 (42.4) | 1,331 (43.2) |  |
| Low-middle | 308 (19.8) | 346 (22.7) | 654 (21.2) |  |
| High-middle | 343 (22.0) | 327 (21.5) | 670 (21.8) |  |
| Highest | 214 (13.8) | 205 (13.4) | 419 (13.6) |  |
| *Missing* | *6 (0.4)* | *n/a* | *6 (0.2)* |  |
| Migration |  |  |  | **<0.001** |
| Long-term resident | 1,119 (71.9) | 1,335 (87.6) | 2,454 (79.7) |  |
| In-migrant | 437 (28.1) | 189 (12.4) | 626 (20.3) |  |
| Community type |  |  |  | **<0.001** |
| Agrarian | 411 (26.4) | 522 (24.2) | 933 (30.3) |  |
| Trading | 339 (21.8) | 312 (20.5) | 651 (21.1) |  |
| Fishing | 806 (51.8) | 690 (45.3) | 1,496 (48.6) |  |
| ***Behavioral*** |  |  |  |  |
| Number of sexual partners |  |  |  | **0.001** |
| 0–1 | 1,099 (70.6) | 1,157 (75.9) | 2,256 (73.2) |  |
| ≥2 | 457 (29.4) | 367 (24.1) | 824 (26.8) |  |
| Condom use |  |  |  | 0.222 |
| No partners or permanent partners only | 1,130 (72.6) | 1,102 (72.3) | 2,232 (72.5) |  |
| Consistent use with casual partners | 216 (13.9) | 189 (12.4) | 405 (13.1) |  |
| Inconsistent use with casual partners | 210 (13.5) | 233 (15.3) | 443 (14.4) |  |
| Transactional sex |  |  |  | 0.178 |
| No | 830 (53.3) | 776 (50.9) | 1,606 (52.1) |  |
| Yes | 726 (46.7) | 748 (49.1) | 1,474 (47.9) |  |
| Any alcohol use consequences |  |  |  | 0.182 |
| No | 1,449 (93.1) | 1,437 (94.3) | 2,886 (93.7) |  |
| Yes | 107 (6.9) | 87 (5.7) | 194 (6.3) |  |
| Illicit drug use |  |  |  | **<0.001** |
| No | 1,464 (94.1) | 1,481 (97.2) | 2,945 (95.6) |  |
| Yes | 92 (5.9) | 43 (2.8) | 135 (4.4) |  |
| ***HIV-related*** |  |  |  |  |
| ART use history (self-reported) |  |  |  | **<0.001** |
| Never | 530 (34.1) | 401 (26.3) | 931 (30.2) |  |
| Currently or previously | 1,026 (65.9) | 1,123 (73.7) | 2,149 (69.8) |  |
| Visit-pair viral load |  |  |  | **<0.001** |
| Durable VLS | 1,006 (64.7) | 1,107 (72.6) | 2,113 (68.6) |  |
| New/renewed VLS | 262 (16.8) | 225 (14.8) | 487 (15.8) |  |
| Viral rebound | 44 (2.8) | 32 (2.1) | 76 (2.5) |  |
| Persistent viremia | 244 (15.7) | 160 (10.5) | 404 (13.1) |  |

**p*-values calculated using Pearson’s chi-square test of association, unless otherwise specified. † *p*-values calculated using Wilcoxon rank-sum tests comparing median values and interquartile ranges (IQR). *^§^* Behavioral factors measured in the past year. *Notes*: VLS was defined using an HIV RNA cutpoint of <200 copies/mL. Demographic variables were derived from the index visit (V*_i_*) in the visit-pair, and behavioral factors were derived from the follow-up visit (V*_i+j_*) in the visit-pair.

**Figure S4.** Box plots of stabilized inverse probability of selection and censoring weights, by number of visit-pairs contributed to the analysis.

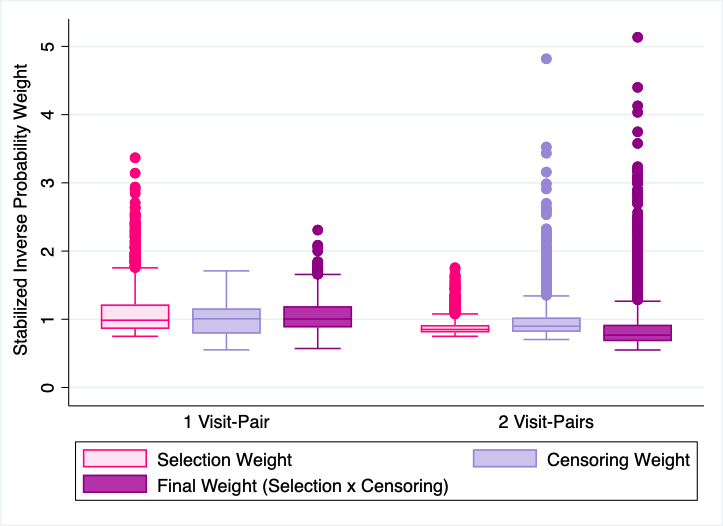

**Table S10.** Quasilikelihood under the independence model criterion (QIC) values for multivariable models estimating the risk of persistent HIV viremia.

| **Working Within-Group**  **Correlation Structure** | **QIC** | **QIC*_u_*** | **Trace** |
| --- | --- | --- | --- |
| Autoregressive | 1731.047 | 1714.686 | 21.181 |
| Exchangeable | 1397.134 | 1404.587 | 9.273 |
| Independent | 1455.061 | 1456.017 | 12.522 |
| Nonstationary | 1731.047 | 1714.686 | 21.181 |
| Stationary | 1731.047 | 1714.686 | 21.181 |
| Unstructured | 1397.134 | 1404.587 | 9.273 |

*Notes*: Multivariable models were estimated using Poisson regression with generalized estimating equations and robust standard errors. QIC*u* represents the penalty attached to the QIC when the working correlation structure is misspecified and is equal to the QIC + 2*p*, where *p* represents the number of parameters estimated in the model.^4^ The trace represents the absolute difference between the QIC*u* and QIC.^5^ The QIC favors working correlation structures yielding smaller values.^5^

**Table S11**. Risk of persistent high-level HIV viremia (≥1,000 copies/mL) relative to sustained or new/renewed low-level viremia or suppression, by gender.

|  | **Men (*N* = 1,146)** | |  | **Women (*N* = 1,882)** | |
| --- | --- | --- | --- | --- | --- |
| **Characteristics** | RR (95% CI) | adjRR (95%CI) |  | RR (95% CI) | adjRR (95% CI) |
| ***Demographics*** |  |  |  |  |  |
| Age group |  |  |  |  |  |
| 15-29 years | **3.07 (2.17–4.35)** | **2.45 (1.71–3.52)** |  | **4.82 (2.87–8.08)** | **3.59 (2.10–6.14)** |
| 30-39 years | **1.49 (1.07–2.07)** | **1.41 (1.02–1.96)** |  | **2.34 (1.42–3.85)** | **2.04 (1.23–3.37)** |
| 40-49 years | *Ref*. | *Ref*. |  | *Ref*. | *Ref*. |
| Currently marital status |  |  |  |  |  |
| Never married | **2.14 (1.50–3.06)** | 1.35 (0.92–1.98) |  | **1.59 (1.02–2.51)** | 1.33 (0.87–2.04) |
| Currently married | *Ref*. | *Ref*. |  | *Ref*. | *Ref*. |
| Previously married | 1.26 (0.94–1.68) | 1.21 (0.92–1.60) |  | 0.83 (0.59–1.17) | 0.88 (0.64–1.23) |
| Completed education |  |  |  |  |  |
| No formal education | 0.77 (0.47–1.27) |  |  | 0.99 (0.53–1.85) | 1.17 (0.63 – 2.18) |
| Primary | *Ref*. |  |  | *Ref*. | *Ref*. |
| Secondary | 0.99 (0.65–1.49) |  |  | **2.21 (1.52–3.22)** | **1.92 (1.34–2.74)** |
| Technical/University | 1.07 (0.42–2.68) |  |  | 0.51 (0.13–2.05) | 0.64 (0.16–2.58) |
| Migration |  |  |  |  |  |
| Long-term resident | *Ref*. | *Ref*. |  | *Ref*. | *Ref*. |
| In-migrant | **1.64 (1.25–2.16)** | 1.21 (0.92–1.58) |  | **1.52 (1.17–1.97)** | 1.11 (0.86–1.43) |
| ***Behavioral*** |  |  |  |  |  |
| Condom use |  |  |  |  |  |
| No partners or permanent partners only | *Ref*. | *Ref*. |  | *Ref*. | *Ref*. |
| Consistent use with casual partners | 1.25 (0.93–1.69) | 1.12 (0.84–1.50) |  | **1.57 (1.52–2.19)** | 1.31 (0.91–1.89) |
| Inconsistent use with casual partners | **1.72 (1.31–2.25)** | **1.38 (1.05–1.80)** |  | 1.30 (0.94–1.80) | 1.15 (0.81–1.63) |
| Transactional sex |  |  |  |  |  |
| No | *Ref*. |  |  | *Ref*. | *Ref*. |
| Yes | 1.00 (0.77–1.30) |  |  | **1.35 (1.04–1.74)** | 1.15 (0.87–1.53) |
| Hazardous alcohol use | **1.11 (1.03–1.19)** | **1.08 (1.01–1.16)** |  | 1.15 (0.99–1.32) | 1.08 (0.95–1.23) |
| Intimate partner violence |  |  |  |  |  |
| None | *Ref*. |  |  | *Ref*. | *Ref*. |
| Any | 1.17 (0.94–1.46) |  |  | **1.36 (1.05–1.75)** | 1.09 (0.84–1.41) |

*Notes*: Risk ratios (RR) and 95% confidence intervals (95%CI) were estimated using Poisson regression with generalized estimating equations, exchangeable covariance matrices, and robust standard errors. Multivariable models were adjusted for survey interval (calendar period) of index visit and all covariates displayed in the columns presenting adjusted results. Sustained or new/renewed low-level viremia or suppression was defined as <1,000 copies/mL across visits or at follow-up only. **Bolded** values represent risk ratios of persistent HIV viremia that were significantly different from the null value of 1 at the *p* < 0.05 level or below. Behavioral factors measured in the past year.

**Table S12**. Risk of persistent high-level HIV viremia (≥1,000 copies/mL) relative to sustained or new/renewed low-level viremia or suppression, by community type.

|  | **Inland Communities (*N* = 1,560)** | |  | **Fishing Communities (*N* = 1,468)** | |
| --- | --- | --- | --- | --- | --- |
| **Characteristics** | RR (95% CI) | adjRR (95% CI) |  | RR (95% CI) | adjRR (95% CI) |
| ***Demographics*** |  |  |  |  |  |
| Age group |  |  |  |  |  |
| 15-29 years | **3.31 (2.30–4.76)** | **3.42 (2.38–4.93)** |  | **3.18 (2.00–5.07)** | **3.00 (1.86–4.86)** |
| 30-39 years | 1.32 (0.92–1.89) | 1.33 (0.92–1.90) |  | **2.23 (1.44–3.44)** | **2.26 (1.46–3.49)** |
| 40-49 years | *Ref*. | *Ref*. |  | *Ref*. | *Ref*. |
| Gender |  |  |  |  |  |
| Man | **1.97 (1.46–2.66)** | **2.16 (1.59–2.94)** |  | **2.19 (1.58–3.04)** | **2.67 (1.94–3.67)** |
| Woman | *Ref.* | *Ref.* |  | *Ref.* | *Ref.* |
| Currently marital status |  |  |  |  |  |
| Never married | 1.44 (0.99–2.11) |  |  | **2.43 (1.53–3.86)** | 1.55 (0.96–2.52) |
| Currently married | *Ref*. |  |  | *Ref*. | *Ref*. |
| Previously married | 0.77 (0.55–1.08) |  |  | 1.20 (0.88–1.64) | 1.21 (0.91–1.62) |
| Completed education |  |  |  |  |  |
| No formal education | 1.06 (0.55–2.05) |  |  | 0.74 (0.45–1.21) | 0.98 (0.60–1.58) |
| Primary | *Ref*. |  |  | *Ref*. | *Ref*. |
| Secondary | 1.37 (0.96–1.96) |  |  | **1.61 (1.05–2.47)** | **1.69 (1.14–2.51)** |
| Technical/University | 0.57 (0.21–1.52) |  |  | 1.87 (0.53–6.59) | 3.10 (0.89–10.79) |
| Household wealth |  |  |  |  |  |
| Lowest | **1.45 (1.00–2.14)** | **1.59 (1.09–2.33)** |  | 0.80 (0.30–1.65) |  |
| Low-middle | **1.60 (1.12–2.29)** | 1.38 (0.97–1.97) |  | 0.52 (0.24–1.17) |  |
| High-middle | **1.52 (1.08–2.12)** | **1.38 (1.01–1.88)** |  | 0.67 (0.29–1.54) |  |
| Highest | *Ref*. | *Ref*. |  | *Ref*. |  |
| Migration |  |  |  |  |  |
| Long-term resident | *Ref*. | *Ref*. |  | *Ref*. | *Ref*. |
| In-migrant | **1.55 (1.17–2.05)** | 1.17 (0.90–1.52) |  | **1.48 (1.13–1.93)** | 1.17 (0.88–1.54) |
| ***Behavioral*** |  |  |  |  |  |
| Number of past-year sexual partners |  |  |  |  |  |
| 0–1 | *Ref*. | *Ref*. |  | *Ref*. | *Ref*. |
| ≥2 | **1.44 (1.10–1.90)** | 0.91 (0.66–1.24) |  | **1.42 (1.07–1.87)** | 0.80 (0.59–1.09) |
| Condom use |  |  |  |  |  |
| No partners or permanent partners only | *Ref*. | *Ref*. |  | *Ref*. | *Ref*. |
| Consistent use with casual partners | **1.60 (1.13–2.27)** | 1.24 (0.87–1.77) |  | **1.44 (1.04–2.00)** | 1.33 (0.92–1.91) |
| Inconsistent use with casual partners | **1.38 (1.01–1.88)** | 1.29 (0.93–1.78) |  | **1.87 (1.38–2.54)** | 1.44 (1.02–2.02) |
| Hazardous alcohol use | **1.25 (1.13–1.37)** | **1.17 (1.08–1.27)** |  | **1.11 (1.02–1.21)** | 1.03 (0.95–1.12) |
| Intimate partner violence |  |  |  |  |  |
| None | *Ref*. |  |  | *Ref*. | *Ref*. |
| Any | 1.15 (0.89–1.48) |  |  | **1.32 (1.05–1.66)** | 1.16 (0.92–1.47) |

*Notes*: Inland communities include all mainland non-fishing (i.e., agrarian and trading) communities. Risk ratios (RR) and 95% confidence intervals (95%CI)

were estimated using Poisson regression with generalized estimating equations, exchangeable covariance matrices, and robust standard errors. Multivariable

models were adjusted for survey interval (calendar period) of index visit and all covariates displayed in the columns presenting adjusted results. Sustained or new/renewed low-level viremia or suppression was defined as <1,000 copies/mL across visits or at follow-up only. **Bolded** values represent risk ratios of persistent HIV viremia that were significantly different from the null value of 1 at the *p* < 0.05 level or below. Behavioral factors measured in the past year.
